# Antenatal Maternal Anaemia and Infant Brain Structure: 3T and 64mT MRI Findings from South Africa

**DOI:** 10.64898/2025.11.28.25341220

**Authors:** Jessica E. Ringshaw, Michal R. Zieff, Niall J. Bourke, Chiara Casella, Layla E. Bradford, Simone R. Williams, Donna Herr, Marlie Miles, Carly Bennallick, Khula South Africa Data Collection Team, Sean Deoni, Jonathan O’Muircheartaigh, Dan J. Stein, Daniel C. Alexander, Derek K. Jones, Steven C.R. Williams, Kirsten A. Donald

**Affiliations:** Department of Paediatrics and Child Health, Red Cross War Memorial Children’s Hospital, University of Cape Town, South Africa; Neuroscience Institute, University of Cape Town, South Africa; Centre for Neuroimaging Sciences, Department of Neuroimaging, Kings College London, England, United Kingdom; Research Department of Early Life Imaging, School of Biomedical Engineering and Imaging Sciences, King’s College London, England, United Kingdom; Department for Forensic and Neurodevelopmental Sciences, Institute of Psychiatry, Psychology and Neuroscience, King’s College London, England, United Kingdom; Maternal, Newborn, Child Nutrition and Health (MNCH) Discovery and Translational (D&T) Sciences Program, Gates Foundation, Seattle, United States of America; Medical Research Council (MRC) Centre for Neurodevelopmental Disorders, King’s College London, England, United Kingdom; South African Medical Research Council (SAMRC), Unit on Risk and Resilience in Mental Disorders, Department of Psychiatry, University of Cape Town, South Africa; Hawkes Institute and Department of Computer Science, University College London, England, United Kingdom; Cardiff University Brain Imaging Research Centre (CUBRIC), School of Psychology, Cardiff University, Wales, United Kingdom

**Keywords:** neuroimaging, high-field mri, ultra-low-field mri antenatal maternal anaemia, child anaemia, iron deficiency, child brain structure

## Abstract

With the evolution of ultra-low-field MRI and the recognition of antenatal maternal anaemia as an important driver of altered neurodevelopment in toddlers and children, it is critical to determine whether these effects are detectable at 64mT in infancy. The aim of this study was to assess the impact of antenatal maternal anaemia on infant brain structure across the first two years of life, using conventional high-field (3T) and ultra-low-field (64mT) MRI.

This neuroimaging sub-study was embedded within Khula, an observational population-based birth cohort in South Africa. Pregnant women were enrolled antenatally and postnatally. Mother-child dyads (*n*=394) were followed prospectively with a subsample attending neuroimaging at approximately 3, 6, 12, 18, and 24 months of age. Anaemia was classified using WHO thresholds and neuroimaging data was processed using MiniMORPH. Linear mixed effects models were used to investigate associations between antenatal maternal anaemia status and absolute regional infant brain volumes using high-field and ultra-low-field MRI.

In repeated measures high-field (*n*=195) and ultra-low-field (*n*=341) infant neuroimaging subsamples, the prevalence of antenatal maternal anaemia was 28.24% (37/131) and 29.76% (61/205), respectively. Maternal anaemia in pregnancy was associated with altered child brain structure across both MRI systems, with group differences becoming detectable at approximately 12 months. In the ultra-low-field subsample, infants born to anaemic mothers had 3.77% smaller ICV (β=-0.24, *p*=0.004) and 3.32% smaller putamen volumes (β=-0.18, *p*=0.04) across the first 2 years of life. The interaction between antenatal maternal anaemia and age was significant for the caudate nucleus (β=-0.13, *p*=0.038) and the corpus callosum (β=-0.13, *p*=0.037). Maternal anaemia in pregnancy was associated with 3.70% and 4.29% smaller caudate nucleus at 18 and 24 months of age, respectively. Similarly, infants born to anaemic mothers had 4.18% smaller corpus callosum volumes by 12 months and 7.10% smaller corpus callosum volumes by 24 months. Postnatal child anaemia and antenatal maternal iron deficiency status were not associated with total or regional child brain volumes in the ultra-low-field subsample from this cohort. Maternal anaemia remained a robust predictor of volume differences in sensitivity analyses.

This study is the first to demonstrate that the impact of maternal anaemia in pregnancy on child brain structure is detectable as early as infancy. The implications of this research are two-fold; (1) informing the feasibility of ultra-low-field MRI for use in low- and middle-income countries, as well as (2) the timing and optimisation of targeted recommendations for anaemia management in both practice and policy.

## INTRODUCTION

Antenatal maternal anaemia, characterised by low serum haemoglobin during pregnancy, affects more than one third (approximately 36% as of 2019) of expecting women worldwide.^1,2^ However, the burden of disease is unevenly distributed with a much higher prevalence observed in low- and middle-income countries (LMICs), particularly in sub-Saharan Africa and South Asia. This is largely driven by risk factors such as malnutrition, infectious diseases, and alcohol use.^3,4^ The impact of the COVID-19 pandemic is likely to have exacerbated this public health problem by disrupting health, educational, and social protection systems while simultaneously increasing the risk of food insecurity.^1,5^ With growing evidence for the foetal origins of brain health and a renewed focus on maternal health in pregnancy,^6^ addressing antenatal maternal anaemia is emerging as a priority for strategies aimed at improving child developmental potential.^7^

While the aetiology of anaemia is multifaceted, chronic iron deficiency is known to be the most common cause due its limiting impact on haemoglobin production.^8^ This negatively affects the haemoglobin-facilitated delivery of oxygenated blood to vital metabolic organs, including the foetal brain during pregnancy.^9,10^ Recent epidemiological research from the Khula South Africa study^11^ has demonstrated that the prevalence of maternal iron deficiency in pregnancy may be as high as 50% after adjustment for inflammation, accounting for half of all anaemia cases in this cohort.^12^ In turn, maternal anaemia and iron deficiency in pregnancy have consistently been identified as risk factors for poor birth^13,14^ and child developmental outcomes,^15,16^ which may have a long-term impact on the next generation’s health, cognition, and behaviour.^1^ This is reiterated by a South African study suggesting that maternal anaemia may be one of the main drivers for children failing to reach their developmental potential.^6^

Thus far, a few Magnetic Resonance Imaging (MRI) studies have suggested that the developing brain may be particularly sensitive to maternal anaemia and iron deficiency in pregnancy,^17–19^ but the results have been largely inconsistent due to widely varying research methods. To address this, a neuroimaging substudy of the Drakenstein Child Health Study (DCHS)^20^ in South Africa compared the relative effects of antenatal maternal and postnatal child anaemia on child brain structure.^7,21^ Maternal anaemia in pregnancy was associated with smaller volumes of the corpus callosum, putamen, and caudate nucleus in toddlers (aged 2-3 years)^7^ and school-age children (aged 6-7 years)^21^ in this cohort. These findings suggest that the effects of antenatal maternal anaemia on child brain structure may be regionally consistent and persist with age, reinforcing the importance of optimizing anaemia interventions in women of reproductive age before and during pregnancy.^7,21^

Despite this emerging evidence, further corroboratory neuroimaging research is required to assess whether the effects of antenatal maternal anaemia on child brain structure can be generalised to other community settings and, critically, whether they can be detected in infancy. Therefore, the primary aim of this study was to investigate the impact of antenatal maternal anaemia on child brain structure in South African children across the first two years of life using conventional high-field (HF; 3T) MRI as proof of concept. The secondary aim of this research was to replicate the HF analyses using ultra-low-field (ULF; 64mT) MRI, a quickly evolving field with the potential to increase neuroimaging accessibility at scale in LMICs. The novel Hyperfine Inc., Swoop® MRI scanner is more cost-effective, more power-efficient, and much less noisy than conventional systems, allowing it to be easily integrated into low-resource clinical settings for paediatric use.^22^ Recent cross-validation research using paired data from the Khula cohort in South Africa has demonstrated high concordance between 3T and 64mT volume estimates for regions of interest known to be associated with maternal anaemia.^23^ While this supports the meaningful interpretation of ULF data in anaemia research, the utility of this system in detecting group differences in regional brain volumes associated with anaemia is largely unknown. Therefore, this research aimed to play a key role in assessing this novel system’s comparative ability to answer clinical questions about antenatal maternal anaemia as detected by conventional MRI in South Africa.

With larger sample sizes and longitudinal postnatal data, the Khula ULF substudy was also well-positioned to delineate the timing of effects by assessing the relative contributions of antenatal maternal anaemia versus postnatal child anaemia on infant brain structure. In parallel, the role of iron deficiency was explored as a potential underlying neurobiological driver of associations between anaemia and regional infant brain volumes. Overall, this research aimed to validate the feasibility of ULF MRI by answering these important clinical questions, while also aiming to inform the timing and optimisation of more targeted recommendations for anaemia management in both practice and policy.

## METHODS

### Study Design and Setting

This research is embedded within Khula South Africa, an observational population-based birth cohort study in Gugulethu, an urban informal settlement in the Western Cape Province of South Africa.^11^ The overarching purpose of this longitudinal study was to characterise executive functioning in the first 1000 days of life, explore the underlying neural pathways in the developing brain, and investigate the role of known risk factors through the use of multi-modal tools including MRI. The aim of this neuroimaging substudy was to use HF (3T) and ULF (64mT) MRI data to investigate the associations between anaemia, iron deficiency, and child brain structure within the first two years of life (3 – 24 months).

Mother-child dyads were recruited antenatally from the Gugulethu Midwife Obstetrics Unit (MOU) and postnatally from clinics in the surrounding area. Previous research from the Khula South Africa cohort^12^ has demonstrated that approximately 65% of mothers are unemployed, with 63% of households earning a total income of less than 5000 ZAR per month (< 300 USD). Additionally, food insecurity was reported by >50% of the mothers and the prevalence of antenatal maternal anaemia was 35%, with approximately half of cases being attributed to iron deficiency after adjustment for inflammation.^12^ Maternal HIV was associated with an increased risk of anaemia in pregnancy, highlighting the relevance of infection as a key aetiological factor for consideration. The demographic and anaemia profile of this cohort is comparable to statistics from the National Department of Health’s (NDH) Demographic and Health Survey (DHS),^24^ making this study community representative of the South African population.

Khula mother-child dyads were followed prospectively with a subsample attending neuroimaging study visits at approximately 3, 6, 12, 18, and 24 months of age.

### Participants

In South Africa, women were recruited (December 2021 – November 2022) if they met the eligibility criteria of being over the age of 18 years and in the third trimester of pregnancy (28-40 weeks’ gestation), or up to 3 months postpartum. Exclusion criteria included multiple pregnancies, psychotropic drug use during pregnancy, infant congenital malformations or abnormalities (e.g., spina bifida, Down’s Syndrome), and severe birth complications (e.g., uterine rupture, birth asphyxia). Overall, a total of 394 mother-child pairs from South Africa were enrolled in the Khula study, with 329 women retained from antenatal recruitment and 65 women retained from postnatal recruitment (Figure 1; Study Flowchart).

**Figure 1.**
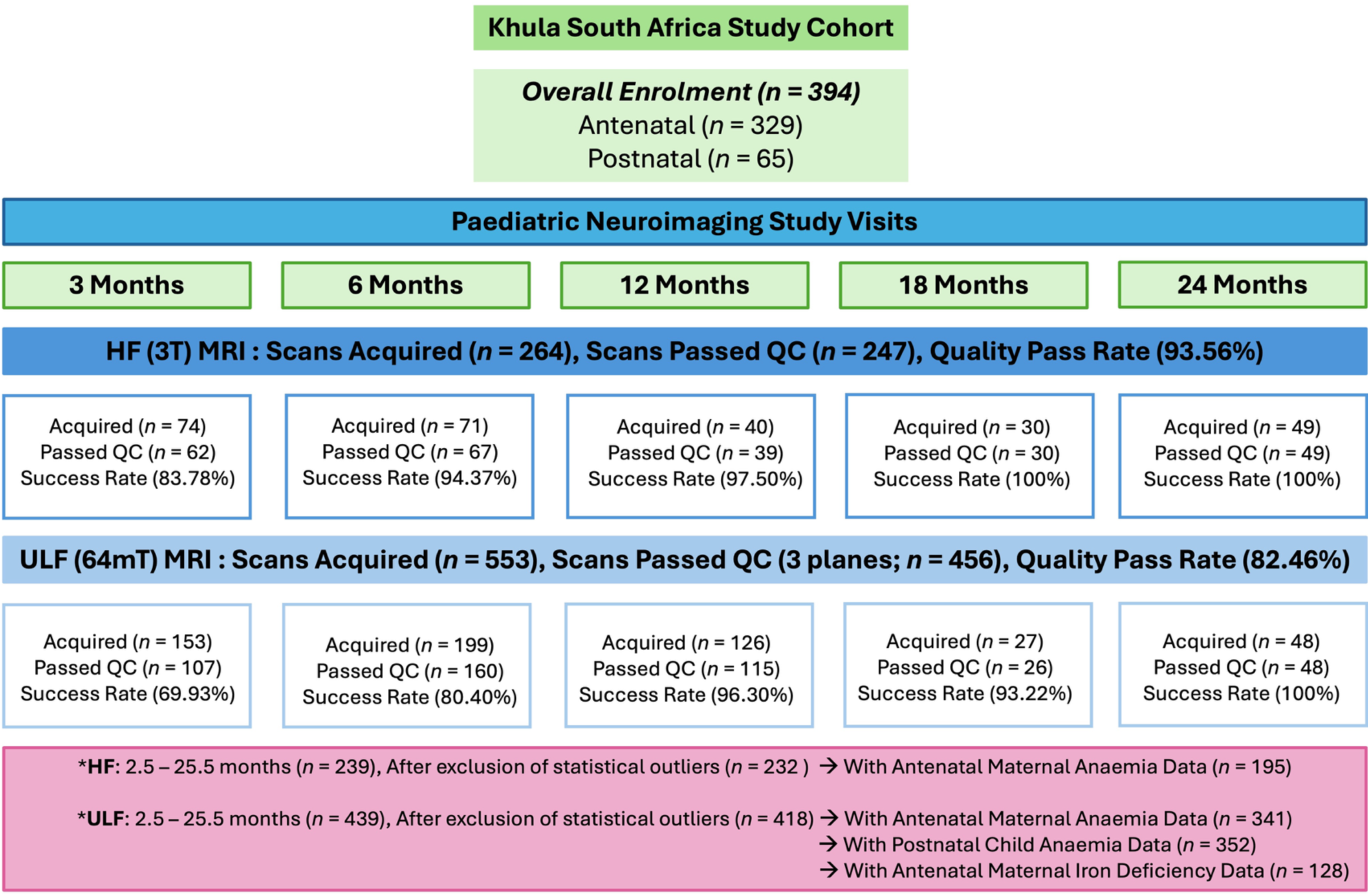
Study Flowchart Demonstrating the Acquisition and Quality Pass Rates of HF (3T) and ULF (64mT) Neuroimaging Data Across Study Visits and The Final Sample Size Determination with Corresponding Antenatal Maternal Anaemia Data The primary analysis assessing the impact of antenatal maternal anaemia on child brain structure was conducted separately using both HF and ULF subsamples. The analyses assessing the role of postnatal child anaemia (secondary analysis) and antenatal maternal iron deficiency (tertiary analysis) on child brain structure were only conducted in the larger ULF subsample due to increased power for the detection of effects.

Of these eligible Khula babies in South Africa, 264 T2-Weighted (T2W) HF scans were acquired across study visits, of which 247 passed quality check (QC) procedures (93.56%). Similarly, 553 ULF T2W scans (complete with axial, coronal, and sagittal planes) were collected across timepoints with 456 (82.46%) passing QC. The neuroimaging subsamples for HF and ULF analyses were compiled for children between 3 and 24 months of age (with a two week window on either side; 2.5 – 25.5 month range) with useable brain scans (passed QC) across timepoints. Statistical outliers were excluded if extreme (*z*-scores above and below 3 standard deviations from the sample mean), or moderate (above and below 2 standard deviations from the sample mean) but consistent across multiple regions of interest (ROIs; 3 for HF to maximise sample size and power; 2 for ULF due to increased risk of measurement error with the ULF system).

Overall, the final neuroimaging subsamples with repeated measures for infants with corresponding antenatal maternal anaemia data included 195 observations for HF scans (131 unique infants) and 341 observations (205 unique infants) for ULF scans (Figure 1; Figure 2). Similarly, the ULF neuroimaging subsample (Figure 1) with repeated measures for infants with corresponding postnatal child anaemia data (secondary analysis) included 352 observations (214 unique infants) and corresponding antenatal maternal iron deficiency (tertiary analysis) included 128 observations (73 unique infants). All sample sizes include repeated measures data available across study timepoints (Figure 2).

**Figure 2.**
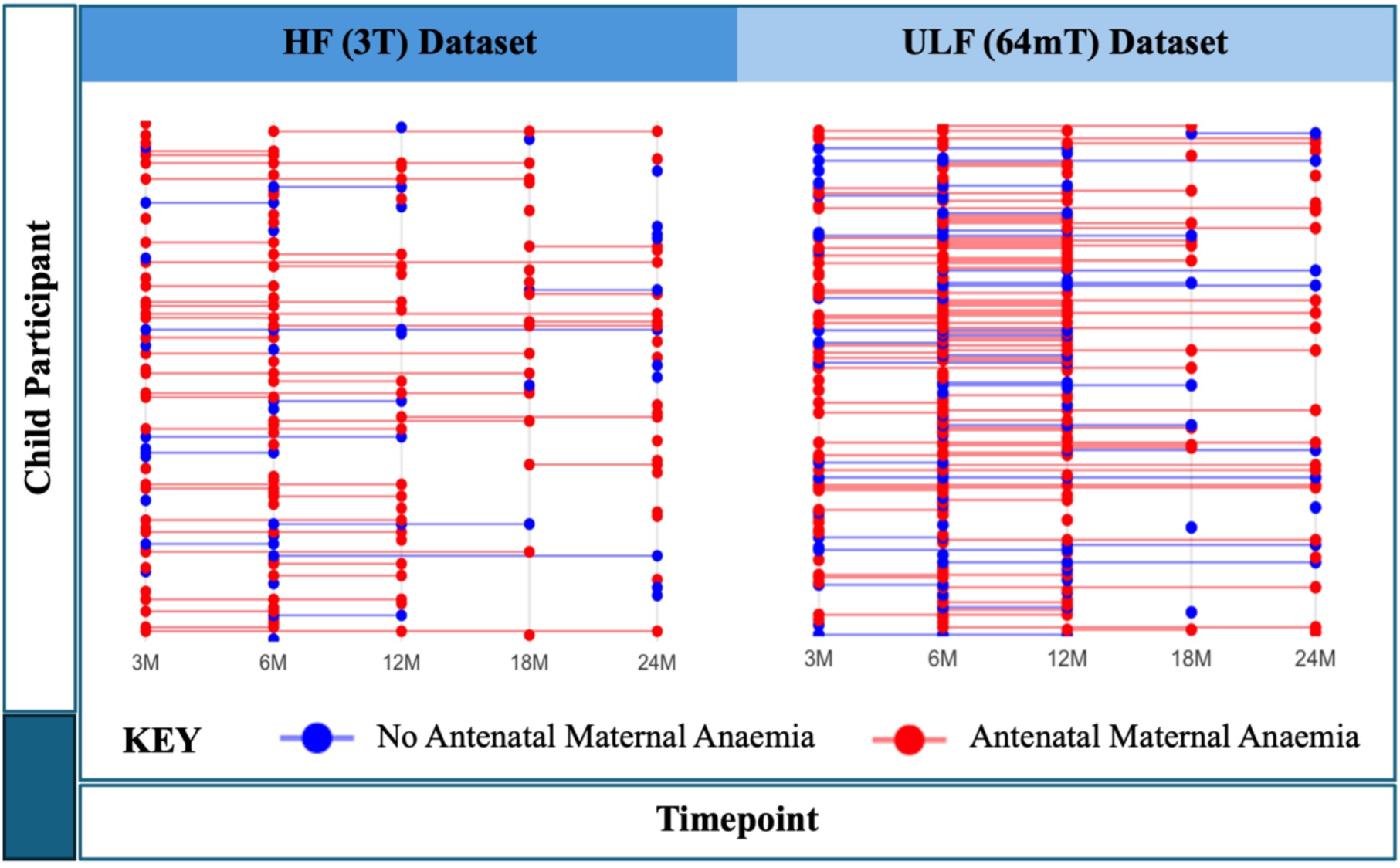
Repeated Measures Data Availability for Participants Across Study Timepoints for HF (3T; Left) and ULF (64mT; Right) Datasets. Timepoints refer to study visits at 3 months (3M), 6 months (6M), 12 months (12M), 18 months (18M), and 24 months (24M). In the HF subsample (*n* = 195 observations; *n* = 131 unique infant subjects), 80/131 (61.10%) of babies only had MRI data available at a single timepoint. Repeated measures data was available for 51/131 (38.93%) babies across 2 timepoints (40/131; 30.5%), 3 timepoints (9/131, 6.9%), and 4 timepoints (2/131; 1.5%). In the ULF subsample (*n* = 341 observations, *n* = 205 unique infant subjects), 101/205 (49.3%) of babies only had MRI data available at a single timepoint. Repeated measures data was available for 104/205 (50.73%) babies across 2 timepoints (75/205; 36.6%), 3 timepoints (26/205; 12.7%), and 4 timepoints (3/205; 1.5%).

### Measures

#### Contextual Measures

Sample characteristics were acquired for demographic (age, sex, socioeconomic status [maternal education, total household income, maternal employment]) and antenatal maternal health (maternal HIV status, maternal depression, prenatal alcohol exposure [PAE], prenatal tobacco exposure [PTE]) variables. This contextual data was previously described for the full cohort^12^ and were included for use as covariates in this neuroimaging substudy.

#### Anaemia Status

Antenatal maternal anaemia status was assessed based on the lowest serum haemoglobin concentration and corresponding gestational age available from a full blood count (FBC) acquired at Khula study enrolment in pregnancy and/or extracted from antenatal medical records (National Health Laboratory Services; NHLS). For mothers with no recorded gestational age data, haemoglobin measurements were assigned to trimester 3 based on likely time of recruitment and highest risk coinciding at this timepoint. Based on the 2024 World Health Organisation (WHO) guidelines, antenatal maternal anaemia status was dichotomously classified using thresholds of <11g/dL in trimesters 1 and 3, and <10.5g/dL in trimester 2.^25^ Further classification based on anaemia severity was defined as mild (haemoglobin 10.0 – 10.9g/dL in trimesters 1 and 3; 9.5 – 10.4g/dL in trimester 2), moderate (haemoglobin 7.0 – 9.9g/dL in trimesters 1 and 3; 7 – 9.4 g/dL in trimester 2), and severe (haemoglobin <7.0g/dL) anaemia.

Haemoglobin was measured for infants at each study visit (3, 6, 12, 18, and 24 months). Dichotomous classifications for postnatal child anaemia were determined for each infant using age-specific cut-offs for haemoglobin based on WHO guidelines^26^ for children over 6 months and local guidelines for children under 6 months. These thresholds have previously been described for this cohort.^12^ An overall classification for child anaemia status was determined based on the diagnosis of anaemia at least once across study visits. This was used for the assessment of group differences in sample characteristics between infants who had ever been anaemic versus infants who had never been anaemic between 3 and 24 months of age. Minimum child haemoglobin and corresponding age across study visits were also determined for each infant and reported on across both groups. For modelling with the repeated measures subsample (multiple scans per child in some instances), child anaemia status was assessed based on the most severe presentation of anaemia as indicated by the minimum haemoglobin measure observed between birth and the time of scan.

### Iron Deficiency Status

Iron deficiency status was assessed using serum ferritin with adjustment for inflammation based on inflammatory biomarkers (highly sensitive C-Reactive Protein [*hs*CRP] and Alpha-1Acid Glycoprotein [AGP]). This was conducted using BRINDA (Biomarkers Reflecting Inflammation and Nutritional Determinants of Anaemia),^27^ a regression modelling approach^28^ that corrects continuous serum ferritin concentrations based on *hs*CRP and AGP concentrations. As previously described in this cohort^12^, iron deficiency status was dichotomously classified using standard WHO thresholds of <12μg/L and <15μg/L for adjusted serum ferritin in infants and women, respectively.^29^

### Neuroimaging Outcomes

#### Scan Acquisition

Both HF and ULF MRI were acquired at the Cape Universities Body Imaging Centre (CUBIC). HF MRI was conducted on a 3T Siemens Skyra scanner using a 16- or 32- channel head coil for infants between 3-12 and 12-18 months of age, respectively. ULF MRI was conducted using the Hyperfine Swoop® Inc., 64mT system. This involved running separate sequences for the acquisition of structural data across axial, coronal, and sagittal planes. Structural T2W sequences were chosen for analysis as this sequence is typically more suitable for assessing brain volume in infants,^30^ and is currently the best-developed sequence on the novel ULF system.^22^ Neuroimaging data were collected during non-sedated sleep using well-established clinical approaches and strategies for paediatric scanning developed by our team.^23,31^ Neuroimaging success rates across field strengths increased with age from the 3-month to 24-month study visit (Figure 1). The neuroimaging protocols and MRI specifications have been described previously.^23^ All HF scans were subject to radiological review and infants with incidental findings were referred via established institutional pathways.

#### Neuroimaging Processing

All HF and ULF scans were subject to a rigorous QC protocol using a standardised checklist for assessing image quality on [Flywheel](https://flywheel.io/), a harmonised data management platform. This was completed by two senior neuroimaging staff members trained to recognise artefacts such as motion and electrical interference. Factors affecting scan success have been reported previously.^23^ Only useable scans that had passed QC (prioritising “good” quality ratings) were included for analyses (Figure 1).

The following processing steps were conducted inside Docker containers via Flywheel ‘gears’ to standardise processing in a reproducible version-controlled manner. The first processing step only applied to the ULF data, and involved using a Multi-Resolution Reconstruction (MRR) pipeline to reconstruct orthogonally-acquired anisotropic images across axial, coronal, and sagittal planes into a single 1.5mm^3^ isotropic image of higher resolution.^32^ The second step of processing used the MiniMORPH pipeline to segment neuroimaging data from across HF and ULF fields strengths.^33^ This is a novel approach using age-specific templates derived from ULF Khula data with specified age-cutoffs for 3, 6, 12, 18, and 24 months. Tissue and CSF priors were generated by aligning white matter, grey matter, and CSF probability maps from the Baby Connectome Project (BCP)^34^ to study templates using SynthMorph.^35^ Tissue priors were created by combining white matter and grey matter maps, and a skull prior was generated by dilating the brain mask for improved classification of non-brain regions. Subcortical and callosal parcellations from the BCP atlas^34^ and the Penn-CHOP Atlas^36^, respectively, were first registered to the 12-month template and then propagated to the other age-specific templates^37^. The corpus callosum was manually subdivided in 5 portions using the Desikan- Killiany atlas.^38^ Ventricle masks were manually delineated in template space. For each scan session in the current study, participants underwent skull stripping with SynthStrip. Tissue priors and masks were registered to native T2-w isotropic images via the age-specific template.^37^ The images were segmented into the 3 tissue classes with bias field correction (tissue, CSF and skull) using ANTs AtroposN4.^37^ The resulting CSF class was multiplied by the ventricle mask to separate CSF from ventricular CSF. The ANTs tissue class was multiplied by the subcortical grey matter and callosal masks to refine subcortical and callosal segmentations. A final native space atlas was completed for each scan with fslmaths including all segmented regions in native space. Volume estimates were extracted with fslstats into csv files for statistical analysis.

### Statistical Analysis

#### Sample Characteristics

Demographic and clinical data were presented as means and standard deviations for continuous variables, and frequencies and percentages for categorical variables. All statistical analyses were conducted in R Studio (2025.05.1+513) for R (version 4.5.1), and a significance level of 0.05 was used throughout. Group differences (based on maternal anaemia status, child anaemia status, and maternal iron deficiency status separately) in sociodemographic and maternal exposure variables were calculated using unpaired *t*-tests for continuous data and chi-squared tests or Fisher’s exact tests for categorical data. This was assessed based on the number of unique subjects in each subsample, excluding repeated measures data for children with multiple scans. This approach was used to avoid skewing results with duplicated demographic and clinical observations for the same infants. The only exception to this approach was child age at scan which was considered for the full repeated measures subsample, with a proportion of children having multiple scans acquired at different ages. Checks for assumptions of normality were conducted throughout.

#### Neuroimaging Regions of Interest

Given that antenatal maternal anaemia has previously been associated with smaller volumes of the caudate nucleus, putamen, and corpus callosum in children at 2-3^7^ and 6-7 years^21^ of age in a similar South African birth cohort, these structures were chosen as key ROIs for a hypothesis-driven analysis. Intracranial volume (ICV) was included to investigate group differences in total brain volume. The total caudate nucleus and total putamen were computed as the sum of left and right hemisphere volumes for these basal ganglia structures. The total corpus callosum was computed as the sum of individually segmented posterior, mid-posterior, central, mid-anterior, and anterior regions.

### Primary Analysis: Antenatal Maternal Anaemia Status

#### Exploratory Analyses

As an exploratory step, the absolute volumes were plotted against child age at the time of scan to visually assess changes in growth with age for ICV and each ROI (putamen, caudate nucleus, corpus callosum). The fit of the data was observed to classify whether regional increases in absolute brain volumes with age represented a linear relationship or a non-linear relationship (locally estimated scatterplot smoothing; LOESS).

#### Statistical Modelling

The associations between antenatal maternal anaemia status and absolute regional infant brain volumes were assessed across field strengths to compare findings for 3T and 64mT MRI systems. The HF dataset was used as proof of concept and to attempt to replicate findings from previously published studies. Analyses were repeated on the ULF system to determine whether the same brain volume trajectories and effects of anaemia could be observed and detected at 64mT.

Given the repeated measures study design with individual infant data across multiple study visits in both HF and ULF subsamples, linear mixed effects (LME) models were fitted to account for individual differences in assessing the association between antenatal maternal anaemia status and absolute regional brain volume trajectories. Child sex at birth and child age at scan were included in all models as covariates known to affect child brain structure *a priori.*^39^ Based on the observed fit of the data (linear vs LOESS) in exploratory analyses, age in months was entered as an absolute value for brain regions with a linear growth trajectory (corpus callosum) or as a logarithmic function for brain regions with an exponential growth trajectory (ICV, putamen, caudate nucleus). ICV was not included in the ROI models due to high levels of demonstrated multicollinearity with age at scan based on zero-order measures correlations in both HF (*r* = 0.814, *p* <0.001) and ULF (*r* = 0.747, *p* < 0.001) subsamples.

Therefore, only scan age was included in models for absolute regional brain volumes, serving as a proxy for ICV. Overall, antenatal maternal anaemia, child age (or the logarithmic function of age), and child sex were entered as fixed effects with a random intercept to account for repeated measures.

Demographic and maternal exposure variables with observed group differences were considered for inclusion in analyses as potential confounders, only if correlated with regional brain volumes in preliminary steps. Based on the plotted data (exploratory analyses), an interaction for the effect of antenatal maternal anaemia status on absolute regional child brain by age (or the logarithmic function of age if relevant for non-linear growth trajectories) was also considered. This was only included as a fixed effect in the ULF models, due to a larger sample size (*n* = 341) and greater statistical power in this dataset. Interactions were excluded from HF analyses due to the smaller sample size (*n* = 195) and the sparse distribution of data across age groups, which may have contributed to unreliable estimates and an increased risk of overfitting.

Post-hoc testing was conducted on the final HF and ULF LMEs using estimated marginal means (EMMs) for the regression line of best fit. For brain regions with no interaction between antenatal maternal anaemia and age, EMMs were used to calculate the overall adjusted percentage difference in regional brain volumes between groups. For brain regions with a significant interaction between antenatal maternal anaemia status and age (or the logarithmic function of age), EMMs were computed for specified age-groups (3, 6, 12, 18, and 24 months). Multiple pairwise comparisons were conducted to determine group differences across ages and to identify the age at which the association between antenatal maternal and region brain volumes became significant. The Holm method was used for multiple comparison correction.^40^

#### Secondary Analysis: Postnatal Child Anaemia Status

The role of postnatal child anaemia was explored using ULF MRI for maximum statistical power. In these LMEs, child anaemia status was based on the most severe presentation of anaemia observed between birth and the time of scan for each repeated measure. First, the exploratory analyses and models described above for maternal anaemia were replicated with postnatal child anaemia as the main predictor of interest. Second, the previously established models assessing the impact of antenatal maternal status on regional brain volumes were re-run with the inclusion of postnatal child anaemia status as a covariate. Finally, a new model was run to establish whether the combination of both antenatal maternal anaemia and postnatal child anaemia was more likely to be associated with smaller infant brain volumes than antenatal maternal anaemia alone.

#### Tertiary Analysis: Iron Deficiency Status

Given that iron deficiency is the most common cause of anaemia, it was investigated as a potential predictor of child brain structure. Based on this proposed aetiological underpinning, it was hypothesized that the association between iron deficiency status and regional brain volumes would mimic that of anaemia status. Therefore, this tertiary analysis was only run for variables with demonstrated associations between anaemia (antenatal maternal versus postnatal child) and child brain structure in primary and secondary analyses; the role of iron deficiency was only assessed if the effects of anaemia were significant.

#### Sensitivity Analyses

Sensitivity analyses for all LME models were conducted to account for the potential confounding role of relevant maternal exposures (with observed group differences in prevalence). Multicollinearity was considered in the establishment of all models.

## RESULTS

### Primary Analysis: Antenatal Maternal Anaemia | HF (3T) and ULF (64mT) MRI

#### Sample Characteristics: Antenatal Maternal Anaemia Status

In the HF neuroimaging subsample (Table 1), 131 mothers (Mean [SD] age at enrolment: 29.79 [5.62] years, age range of 19.00 – 41.70 years) had antenatal maternal haemoglobin data available. Minimum haemoglobin measurements were predominantly acquired in the second (35.88% [47/131]) and third trimester of pregnancy (34.35% [45/131], at a median (IQR) of 21.50 (14 – 32) weeks. Of these mothers, 28.24% (37/131) were classified as anaemic during pregnancy, with 70.27% (26/37) of these cases being mild, 27.03% (10/37) moderate, and 2.70% (1/37) severe. With repeated measures available for 38.93% (51/131) of the subsample included (Figure 2), a total of 195 infant scans (average age at time of scan: 11.91 [6.37] months, age range of 2.56 – 24.84 months) with corresponding maternal anaemia data were available. There were no differences in maternal characteristics or child characteristics between groups based on antenatal maternal anaemia status, with the exception of household income (*p* = 0.031) which was lower in mothers with anaemia than non-anaemic mothers.

**Table 1.** Maternal and Infant Sample Characteristics According to Antenatal Maternal Anaemia Status across HF (3T) and ULF (64mT) Subsamples.

| Variable<sup>a</sup> | HF (3T) Subsample (*n* = 131)<sup>a</sup> |  |  | ULF (64mT) Subsample (*n* = 205)<sup>a</sup> |  |  |
| --- | --- | --- | --- | --- | --- | --- |
|  | Maternal Anaemia<br>(*n* = 37) | No Maternal Anaemia<br>(*n* = 94) | *p* | Maternal Anaemia<br>(*n* = 61) | No Maternal Anaemia<br>(*n* = 144) | *p* |
| **Antenatal Maternal Anaemia Profile <sup>b</sup>** |  |  |  |  |  |  |
| Severity in pregnancy |  |  |  |  |  |  |
| Mild | 26 (70.27) | N/A | N/A | 41 (67.21) | N/A | N/A |
| Moderate | 10 (27.03) | N/A | N/A | 19 (31.15) | N/A | N/A |
| Severe | 1 (2.70) | N/A | N/A | 1 (1.64) | N/A | N/A |
| Gestational age at Hb measurement | 27.39 (8.22)<br>[7.00 – 40.00] | 20.95 (10.40)<br>[5.00 – 41.00] | 0.002** | 26.21 (9.48)<br>[6.00 – 39.00] | 21.07 (9.90)<br>[5.00 – 41.00] | <0.001*** |
| Pregnancy trimester Hb measured <sup>g</sup> |  |  |  |  |  |  |
| First | 2 (5.41) | 25 (26.60) | 0.011* | 9 (14.75) | 33 (22.92) | 0.010* |
| Second | 13 (35.14) | 34 (36.17) |  | 18 (29.51) | 59 (40.97) |  |
| Third | 8 (21.62) | 27 (28.72) |  | 29 (47.54) | 37 (25.69) |  |
| Trimester Unknown | 14 (37.84) | 8 (8.51) |  | 5 (8.20) | 15 (10.42) |  |
| Minimum Hb in pregnancy (g/dL) |  |  |  |  |  |  |
| Trimester 1 (HF: *n* = 27, ULF: *n* = 42) | 10.45 (0.64)<br>[10.00 – 10.90] | 12.18 (0.70)<br>[11.00 – 13.50] | 0.002** | 10.26 (0.96)<br>[7.90 – 10.90] | 12.12 (0.69)<br>[11.00 – 13.50] | <0.001*** |
| Trimester 2 (HF: *n* = 47, ULF: *n* = 77) | 9.78 (0.45)<br>[8.80 – 10.30] | 11.82 (0.84)<br>[10.60 – 13.80] | <0.001*** | 9.77 (0.65)<br>[7.7 – 10.30] | 11.68 (0.72)<br>[10.60 – 13.20] | <0.001*** |
| Trimester 3 (HF: *n* = 35, ULF: *n* = 66 ) | 9.54 (1.39)<br>[5.30 – 10.90] | 11.65 (0.62)<br>[11.00 – 13.50] | <0.001*** | 9.61 (1.30)<br>[4.40 – 10.90] | 11.64 (0.61)<br>[11.00 – 13.50] | <0.001*** |
| **Maternal Characteristics** |  |  |  |  |  |  |
| Monthly household income (ZAR) <sup>g,g</sup> |  |  |  |  |  |  |
| <1000 | 8 (21.62) | 16 (17.02) | 0.031* | 12 (19.67) | 24 (16.67) | 0.022* |
| 1000-5000 | 24 (64.86) | 36 (38.30) |  | 35 (57.38) | 54 (37.50) |  |
| 5000-10000 | 5 (13.51) | 24 (25.53) |  | 11 (18.03) | 38 (26.39) |  |
| >10000 | 0 (0.00) | 8 (8.51) |  | 0 (0.00) | 10 (6.94) |  |
| Education <sup>f</sup> |  |  |  |  |  |  |
| Primary | 1 (2.70) | 3 (3.19) | 0.405 | 1 (1.64) | 2 (1.39) | 0.362 |
| Some secondary | 21 (56.76) | 40 (42.55) |  | 34 (55.74) | 59 (40.97) |  |
| Completed secondary | 12 (32.43) | 30 (31.91) |  | 20 (32.79) | 60 (41.67) |  |
| Some tertiary | 2 (5.41) | 11 (11.70) |  | 4 (6.56) | 13 (9.03) |  |
| Completed tertiary | 1 (2.70) | 10 (10.64) |  | 2 (3.28) | 10 (6.94) |  |
| Employed <sup>g</sup> | 12 (32.43) | 32 (34.04) | 0.992 | 21 (34.43) | 50 (34.72) | 1 |
| Age at enrolment (years) | 29.75 (6.18)<br>[20.7 – 40.5] | 29.80 (5.42)<br>[19.0 – 41.7] | 0.965 | 29.18 (5.58)<br>[18.5 – 40.3] | 28.87 (5.75)<br>[18.00 – 41.7] | 0.723 |
| Smoking during pregnancy <sup>f</sup> | 1 (2.70) | 3 (3.19) | 1 | 4 (6.56) | 3 (2.08) | 0.200 |
| Alcohol during pregnancy <sup>f</sup> | 3 (8.11) | 3 (3.19) | 0.350 | 4 (6.56) | 7 (4.86) | 0.736 |
| Depression during pregnancy | 4 (10.81) | 18 (19.15) | 0.374 | 9 (14.75) | 25 (17.36) | 0.800 |
| HIV infection during pregnancy | 18 (48.65) | 28 (29.79) | 0.067 | 30 (49.18) | 39 (27.08) | 0.004** |

| **Infant Characteristics** |  |  |  |  |  |  |
| --- | --- | --- | --- | --- | --- | --- |
| Sex (male) | 17 (45.95) | 48 (51.06) | 0.739 | 24 (39.34) | 74 (51.39) | 0.154 |
| Gestational age at birth (weeks) | 39.00 (2.08) | 39.30 (1.72) | 0.419 | 38.88 (2.02) | 39.23 (1.80) | 0.233 |
|  | [33 – 42] | [32 – 42] |  | [32.00 – 42.00] | [30.00 – 42.00] |  |
| Child age at scan (months; $n = 341$)<sup>h</sup> | 12.33 (6.78) | 11.77 (6.23) | 0.590 | 10.64 (5.43) | 11.13 (5.38) | 0.446 |
|  | [2.66 – 24.84] | [2.56 – 24.11] |  | [2.66 – 25.49] | [2.69 – 25.07] |  |
| HIV infection<sup>g</sup> | 0 (0.00) | 0 (0.00) | N/A | 1 (1.64) | 0 (0.00) | 0.436 |
| Birth weight (g) | 3121.19 (486.63) | 3239.19 (513.51) | 0.233 | 3079.75 (471.40) | 3191.30 (511.11) | 0.146 |
|  | [1975.00 – 4360.00] | [1810.00 – 4750.00] |  | [1975.00 – 4360.00] | [1180.00 – 4750.00] |  |
| Birth length (cm) | 49.61 (3.83) | 50.26 (3.12) | 0.296 | 49.34 (3.44) | 50.08 (3.54) | 0.177 |
|  | [41.00 – 57.00] | [43.00 – 57.00] |  | [41.00 – 57.00] | [33.00 – 57.00] |  |
| Birth head circumference (cm) | 34.54 (1.69) | 34.82 (1.80) | 0.417 | 34.38 (1.60) | 34.55 (1.81) | 0.518 |
|  | [30.50 – 38.00] | [31.00 – 40.00] |  | [30.50 – 38.00] | [28.00 – 40.00] |  |

Abbreviations: Hb, haemoglobin; ZAR, South African Rand; HIV, Human Immunodeficiency Virus; COVID-19, Coronavirus Disease 2019; g, grams; cm, centimetres.
<sup>a</sup> The total and group subsamples represent the number of unique subjects, excluding repeated measures data for children with multiple scans, to avoid duplication of observations for infants with multiple scans. Values for continuous variables are presented as: mean (standard deviation) [range]. Values for categorical variables are presented as: count (%).
<sup>b</sup> Maternal anaemia during pregnancy was classified according to the WHO threshold of <11g/dL for trimesters 1 and 3, and <10.5g/dL for trimester 2. Severity categorisations were made indicating mild (haemoglobin 10.0 – 10.9g/dL in trimester 1 and 3; 9.5-10.4 in trimester 2), moderate (haemoglobin 7.0 – 9.9g/dL in trimester 1; 7-9.4 in trimester 2), and severe (haemoglobin <7.0g/dL) anaemia categories.
<sup>c</sup> Trimester of pregnancy defined as first (0-12 weeks), second (13-27 weeks), and third (28 weeks onwards).
<sup>d</sup> The birth anthropometry were conducted by trained labour staff in the ward. Infant length was measured in cm to the nearest completed 0.5cm and weight was measured in kgs.
<sup>e</sup> Levene's test was significant. *T*-test results were interpreted based on equal variance not assumed (Welch two-sample *t*-test).
<sup>f</sup> Fisher's exact test result interpreted due to one or more cells having an expected count of less than 5.
<sup>g</sup> Missing values: pregnancy trimester Hb measured (HF: $n = 12$ , ULF: $n = 20$ ), gestational age at Hb measurement (HF: $n = 12$ , ULF: $n = 20$ ), monthly household income (HF: $n = 10$ , ULF: $n = 21$ ), maternal employment (HF: $n = 1$ , ULF: $n = 1$ ), infant HIV infection (HF: $n = 89$ , ULF: 143)
<sup>h</sup> Mean child age at scan was calculated for the full subsample of scans ( $n = 341$ ) including repeated measures, given that each child may have had multiple scans at different ages from different study visits. The group summary statistics for child age at scan represent the mean, standard deviation, and range for each independent observation.
\* $p$ is significant at <0.05, \*\* $p$ is significant at <0.01, \*\*\* $p$ is significant at <0.001.

In the ULF infant neuroimaging subsample (Table 1), 205 mothers (Mean [SD] age at enrolment: 28.96 [5.69] years, age range of 18.00 – 41.70 years) had antenatal maternal haemoglobin data available. Minimum haemoglobin measurements were mostly acquired in the second (37.56% [77/205]) and third trimester of pregnancy (32.20% [66/205], at a median (IQR) of 22 (13 – 30) weeks. Of these mothers, 29.76% (61/205) were classified as anaemic during pregnancy, with 67.21% (41/61) of these cases being mild, 31.15% (19/61) moderate, and 1.64% (1/67) severe. With repeated measures available for 50.73% (104/205) of the subsample included (Figure 2), a total of 341 infant scans (average age at time of scan: 10.98 [5.39] months, age range of 2.66 – 25.49 months) with corresponding maternal anaemia data were available. There were no differences in maternal characteristics or child characteristics between groups based on antenatal maternal anaemia status (*n* = 205), with the exception of household income (*p* = 0.022) and antenatal maternal HIV (*p* = 0.004). Mothers with anaemia in pregnancy were found to have lower household income and a higher prevalence of HIV infection (49.18% versus 27.08%) than mothers without anaemia in pregnancy.

#### Exploratory Analyses: Antenatal Maternal Anaemia Status

In the full repeated measures subsamples of infants with HF (*n* = 195) and ULF (*n* = 341) neuroimaging data and corresponding antenatal maternal haemoglobin data, absolute brain volumes for ICV and ROIs were plotted (Figure 3) against age to determine the developmental patterns of growth from 3 – 24 months. Overall, for both HF and ULF subsamples, ICV and the basal ganglia structures (caudate nucleus and putamen) fit a loess curve with exponential growth in early life that plateaued after 12 months of age. In contrast, the corpus callosum demonstrated a linear relationship with age, steadily increasing in absolute volume across the first two years of life.

**Figure 3.**
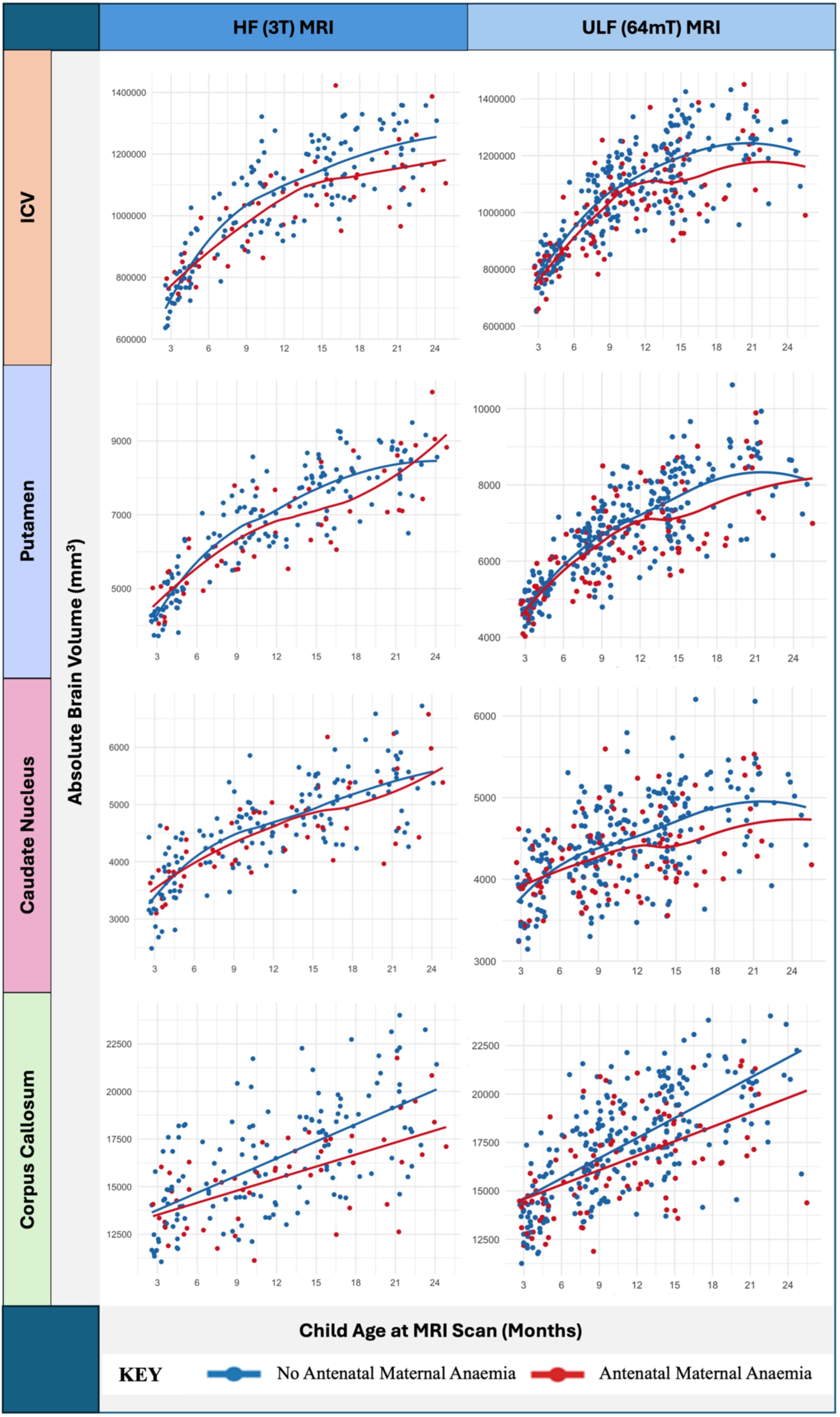
HF (3T; Left) and ULF (64mT; right) Plots of Total and Absolute Regional Infant Brain Volumes By Age and Antenatal Maternal Anaemia Status

This general trajectory for ICV and absolute basal ganglia volumes was apparent in the HF subsample for both groups, with consistently lower volumes in children whose mothers were anaemic in pregnancy (Figure 3). However, in the larger ULF subsample, a more prominent difference between groups was observed at 12 months, with lower ICV and basal ganglia volumes in infants of anaemic versus non-anaemic mothers thereafter. In contrast, the corpus callosum volume increased at a slower rate in infants whose mothers were anaemic during pregnancy than infants whose mothers were not. This contributed to the observed difference between corpus callosum volumes across groups becoming increasingly more prominent across this age group (Figure 3).

#### Modelling: Antenatal Maternal Anaemia Status

Following on from the exploratory plots (Figure 3), zero-order correlation matrices were compiled to preliminarily assess relationships between variables of interest for both HF (*n* = 195) and ULF (*n* = 341) datasets. Thereafter, LME models were fitted to assess the association between antenatal maternal anaemia status and absolute regional brain volumes, with a random intercept to account for repeated measures. Based on the visually inferred linear relationship between child age and absolute total corpus callosum volumes (Figure 3), all LMEs for this region were run with absolute age in months. In contrast, the logarithmic function of age was used for HF and ULF models of brain regions with non-linear growth trajectories (ICV, putamen, and caudate nucleus). Given that child age and child sex were correlated with all brain regions of interest, and considering their known effect on brain structure,^39^ they were included as covariates in both HF and ULF models. Only main effects were tested for the HF subsample, while interactions between antenatal maternal anemia status and age were explored for the larger ULF subsample. All models were was conducted separately for each brain region of interest.

Additional potentially confounding variables with observed differences in frequency between maternal anaemia groups (Table 1) were also considered for inclusion in the models based on zero-order correlations (Supplementary Material; Table S1). In the HF repeated measures subsample (*n* = 195), household income was lower in anaemic mothers (Table 1) but was not independently associated with total or regional brain volumes. In the ULF repeated measures subsample (*n* = 341), lower household income and maternal HIV (Table 1) were significantly correlated with each other and with antenatal maternal anaemia status. This suggests that women living with HIV in lower income households may be at increased risk of being anaemic. However, neither were independently associated with total or regional brain volumes of interest. Therefore, to avoid multicollinearity, these covariates were excluded from HF and ULF models.

#### ICV

While no association between antenatal maternal anaemia and ICV was observed in the HF model (Table 2; *n* = 195; β = -0.15, *p* = 0.111), a significant main effect did emerge when assessed in a bigger subsample at ULF (Table 3; *n* = 341; β = -0.24, *p* = 0.004). Analysis of EMMs in the ULF subsample indicated that infants born to mothers with anaemia (EMM = 1030186.00mm^3^, SE = 11643.87mm^3^) had 3.77% smaller ICV than infants born to mothers without (EMM = 1070599.00mm^3^, SE = 7525.96mm^3^). No interaction was observed between antenatal maternal anaemia and child age at ULF scan (β = -0.04, *p* = 0.311). However, the divergence between group ICV volume trajectories appeared more apparent at 12 months of age, suggesting that the effect of antenatal maternal anaemia on ICV may be more easily detectable in the second year of life (Figure 3).

**Table 2.** Estimated Marginal Means and Adjusted Volume Differences for Models in the HF Repeated Measures Subsample (*n* = 195)

| Brain Region | Estimated Marginal Mean (EMM) Volume ($\text{mm}^3$)<sup>a,b</sup> | | | | $p$ |
| --- | --- | --- | --- | --- | --- |
| | Maternal Anaemia ($n = 51$)<br>EMM (SE) | No Maternal Anaemia ($n = 144$)<br>EMM (SE) | Adjusted Mean Difference (95% CI) | Adjusted Mean Difference (%) | |
| ICV | 1015568.00<br>(14274.92) | 1042549.00<br>(8899.98) | -26981.00<br>(-60277.00 – 6315.00) | -2.59 | 0.111 |
| Putamen | 6644.00<br>(112.00) | 6771.00<br>(69.6) | -127.00<br>(-388.00 – 133.00) | -1.88 | 0.336 |
| Caudate Nucleus | 4540.91<br>(84.30) | 4577.23<br>(52.46) | -36.32<br>(-233.00 – 160.00) | -0.79 | 0.715 |
| Corpus Callosum | 15594.77 (351.67) | 16449.43 (218.51) | -854.66<br>(-1674.00 – 35.2) | -5.20 | 0.041* |

<sup>a</sup> Linear mixed effects (LME) models were fitted for HF data with antenatal maternal anaemia status, child age at scan, child sex, and a random intercept to account for repeated measures. Absolute child age in months was entered for brain regions with linear growth (corpus callosum) and log of age was entered for brain regions with non-linear growth (ICV, putamen, caudate nucleus). No interactions were examined for HF model fitting due to limited power and sparse sample distribution across age-groups.
<sup>b</sup> EMMs are based on the regression line for fully adjusted models. These means are estimated based on the fit of the data, taking into account the role of other covariates.
\* $p$ is significant at $<0.05$ , \*\* $p$ is significant at $<0.01$ , \*\*\* $p$ is significant at $<0.001$

**Table 3.**
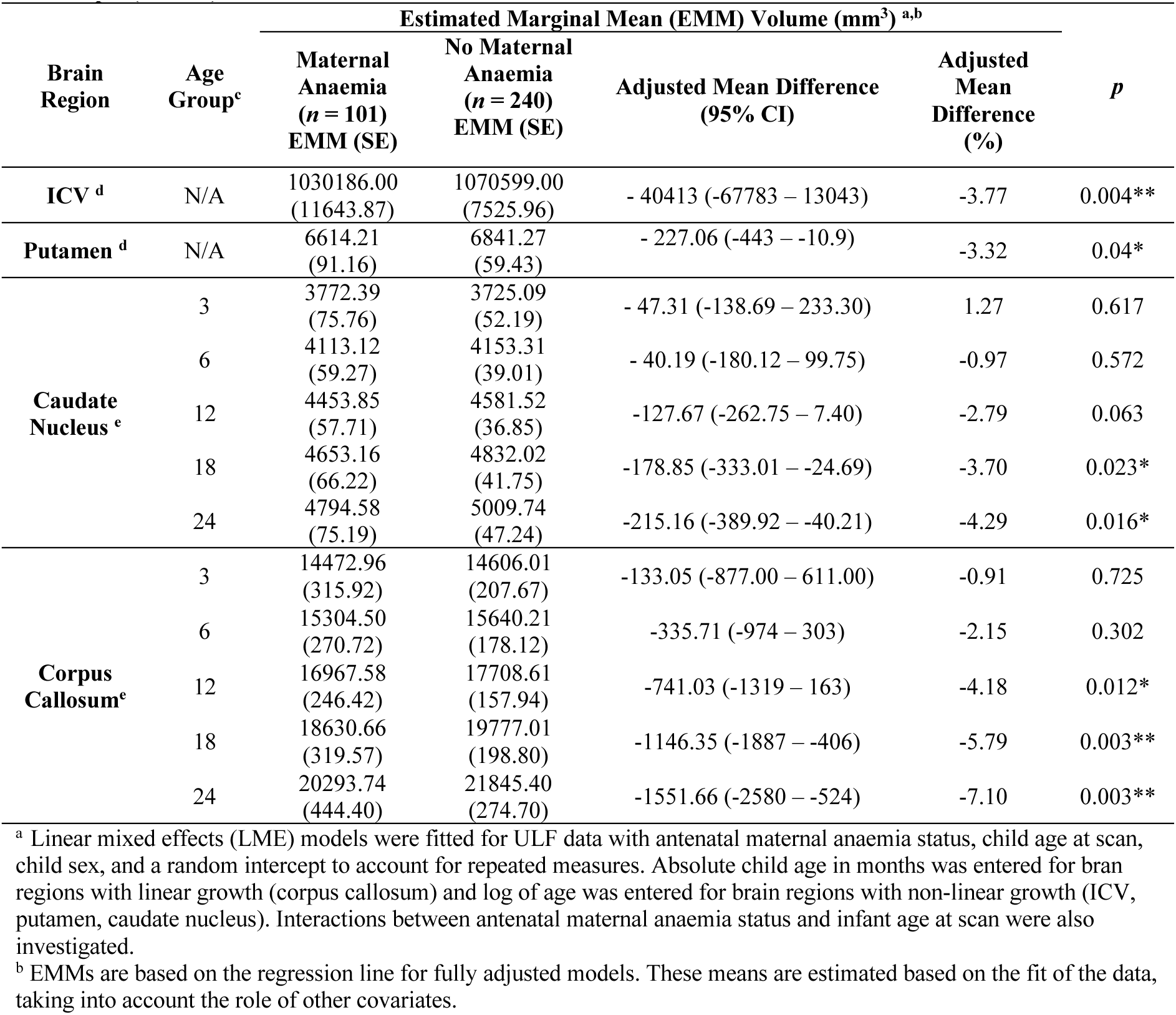
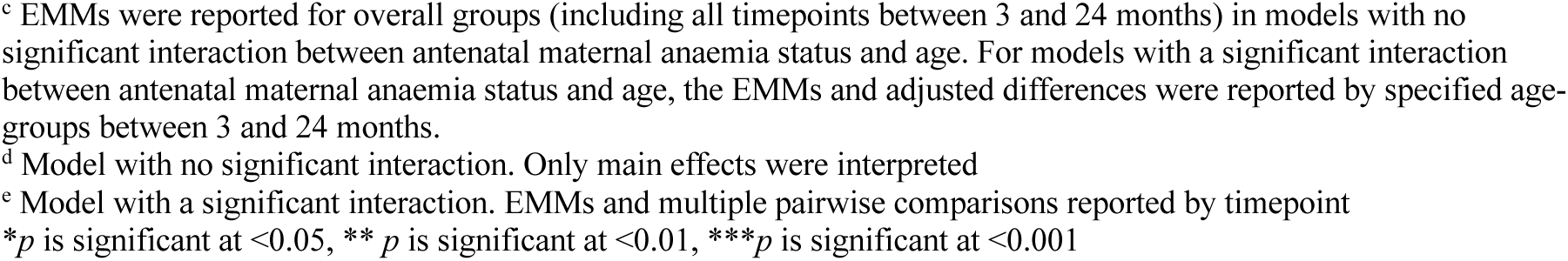
Estimated Marginal Means and Adjusted Differences for Final Models in the ULF Repeated Measures Subsample (*n* = 341)

| Brain Region | Age Group<sup>c</sup> | Estimated Marginal Mean (EMM) Volume ($\text{mm}^3$)<sup>a,b</sup> | | | | $p$ |
| --- | --- | --- | --- | --- | --- | --- |
| | | Maternal Anaemia ($n = 101$)<br>EMM (SE) | No Maternal Anaemia ($n = 240$)<br>EMM (SE) | Adjusted Mean Difference (95% CI) | Adjusted Mean Difference (%) | |
| ICV<sup>d</sup> | N/A | 1030186.00<br>(11643.87) | 1070599.00<br>(7525.96) | - 40413 (-67783 – 13043) | -3.77 | 0.004** |
| Putamen<sup>d</sup> | N/A | 6614.21<br>(91.16) | 6841.27<br>(59.43) | - 227.06 (-443 – -10.9) | -3.32 | 0.04* |
| Caudate Nucleus<sup>e</sup> | 3 | 3772.39<br>(75.76) | 3725.09<br>(52.19) | - 47.31 (-138.69 – 233.30) | 1.27 | 0.617 |
|  | 6 | 4113.12<br>(59.27) | 4153.31<br>(39.01) | - 40.19 (-180.12 – 99.75) | -0.97 | 0.572 |
|  | 12 | 4453.85<br>(57.71) | 4581.52<br>(36.85) | -127.67 (-262.75 – 7.40) | -2.79 | 0.063 |
|  | 18 | 4653.16<br>(66.22) | 4832.02<br>(41.75) | -178.85 (-333.01 – -24.69) | -3.70 | 0.023* |
|  | 24 | 4794.58<br>(75.19) | 5009.74<br>(47.24) | -215.16 (-389.92 – -40.21) | -4.29 | 0.016* |
| Corpus Callosum<sup>e</sup> | 3 | 14472.96<br>(315.92) | 14606.01<br>(207.67) | -133.05 (-877.00 – 611.00) | -0.91 | 0.725 |
|  | 6 | 15304.50<br>(270.72) | 15640.21<br>(178.12) | -335.71 (-974 – 303) | -2.15 | 0.302 |
|  | 12 | 16967.58<br>(246.42) | 17708.61<br>(157.94) | -741.03 (-1319 – 163) | -4.18 | 0.012* |
|  | 18 | 18630.66<br>(319.57) | 19777.01<br>(198.80) | -1146.35 (-1887 – -406) | -5.79 | 0.003** |
|  | 24 | 20293.74<br>(444.40) | 21845.40<br>(274.70) | -1551.66 (-2580 – -524) | -7.10 | 0.003** |

<sup>a</sup> Linear mixed effects (LME) models were fitted for ULF data with antenatal maternal anaemia status, child age at scan, child sex, and a random intercept to account for repeated measures. Absolute child age in months was entered for brain regions with linear growth (corpus callosum) and log of age was entered for brain regions with non-linear growth (ICV, putamen, caudate nucleus). Interactions between antenatal maternal anaemia status and infant age at scan were also investigated.
<sup>b</sup> EMMs are based on the regression line for fully adjusted models. These means are estimated based on the fit of the data, taking into account the role of other covariates.
<sup>c</sup> EMMs were reported for overall groups (including all timepoints between 3 and 24 months) in models with no significant interaction between antenatal maternal anaemia status and age. For models with a significant interaction between antenatal maternal anaemia status and age, the EMMs and adjusted differences were reported by specified age-groups between 3 and 24 months.
<sup>d</sup> Model with no significant interaction. Only main effects were interpreted
<sup>e</sup> Model with a significant interaction. EMMs and multiple pairwise comparisons reported by timepoint
\* $p$ is significant at $<0.05$ , \*\* $p$ is significant at $<0.01$ , \*\*\* $p$ is significant at $<0.001$

Child age (HF: β = 0.80, *p* < 0.001; ULF: β = 0.80, *p* < 0.001) and child sex (HF: β = 0.40, *p* < 0.001; ULF: β = 0.42, *p* < 0.001) were also significant predictors in both HF and ULF models, with larger ICV observed in boys and with increasing age.

#### Corpus Callosum

In the primary LME model for the HF subsample (Table 2; *n* = 195), antenatal maternal anaemia was associated with corpus callosum volumes (*p* = β = - 0.30, 0.0411). Analysis of EMMs indicated that infants born to mothers with anaemia (EMM = 15594. 77mm^3^, SE = 351.67mm^3^) had 5.20% smaller volumes of the corpus callosum than infants born to mothers without (EMM = 16449.43mm^3^, SE = 218.51mm^3^). Building on from this, an interaction between age and antenatal maternal anaemia status was detected with ULF (Table 3; *n* = 341; β = -0.13, *p* = 0.037), indicating that the impact of antenatal maternal anaemia status on corpus callosum volumes may vary in magnitude with age. Analysis of post-hoc EMMs revealed that group differences in corpus callous volumes only became evident at 12 months of age (*p* = 0.012). In this age group, children born to mothers with anaemia in pregnancy (EMM: 16967.58 mm^3^, SE = 246.42mm^3^) had 4.18% smaller corpus callosum volumes than children born to mothers without (EMM = 17708.61mm^3^, SE = 157.94mm^3^). However, by 24 months of age, children born to mothers with anaemia in pregnancy (EMM = 20293.74mm^3^, SE = 444.40mm^3^) had 7.10% smaller corpus callosum volumes than children born to mothers without (EMM = 21845.40mm^3^, SE = 274.70mm^3^). Overall, the difference in corpus callosum volumes between groups became more prominent with age in the first 2 years of life (Figure 3).

Child sex (HF: β = 0.34, *p* = 0.011; ULF: β = 0.45, *p* < 0.001) and child age (HF: β = 0.56, *p* < 0.001; ULF: β = 0.69, *p* < 0.001) were also significant predictors in the model, suggesting that increasing age and being biologically male were associated with larger corpus callosum volumes in infants between 3 and 24 months of age, both expected findings.

#### Caudate Nucleus and Putamen

In the primary model for the HF subsample (Table 2; *n* = 195), antenatal maternal anaemia was not associated with caudate nucleus (β = -0.04, *p* = 0.715) or putamen volumes (β = -0.08, *p* = 0.336). However, in the ULF subsample (Table 3; *n* = 341), antenatal maternal anaemia was negatively associated with putamen volumes (β = -0.18, *p* = 0.04). Analysis of EMMs in the ULF subsample indicated that infants born to mothers with anaemia (EMM = 6614.21mm^3^, SE = 91.96mm^3^) had 3.32% smaller putamen than infants born to non-anaemic mothers (EMM = 6841.27mm^3^, SE = 59.43mm^3^). No interaction was observed between antenatal maternal anaemia and child age for the putamen at ULF (β = -0.03, *p* = 0.519). In contrast, the caudate nucleus was associated with a significant interaction between age and antenatal maternal anaemia status (β = -0.13, *p* =0.038). This suggests that the impact of antenatal maternal anaemia status on caudate nucleus volumes may vary in magnitude with age, with between-group differences emerging after 12 months of age (Figure 3). Analysis of post-hoc EMMs revealed that group differences in caudate nucleus volumes only became evident at 18 months of age (*p* = 0.023). In this age group, children born to mothers with anaemia in pregnancy (EMM: 4653.16mm^3^, SE = 66.22mm^3^) had 3.70% smaller caudate nucleus volumes than children born to mothers without (EMM = 4832.02mm^3^, SE = 41.75mm^3^). By 24 months of age, children born to mothers with anaemia in pregnancy (EMM = 4794.58mm^3^, SE = 75.19mm^3^) had 4.29% smaller caudate nucleus volumes than children born to mothers without (EMM = 5009.74mm^3^, SE = 47.24mm^3^).

As expected, child age (HF: β = 0.86, *p* < 0.001; ULF: β = 0.83, *p* < 0.001) and child sex (HF: β = 0.29, *p* < 0.001; ULF: β = 0.35, *p* < 0.001) were positively associated with putamen volumes in both HF and ULF models. This relationship for child age (HF: β = 0.72, *p* < 0.001; ULF: β = 0.63, *p* < 0.001) and sex (HF: β = 0.30, *p* = 0.007; ULF: β = 0.46, *p* < 0.001) was also observed for the caudate nucleus. As observed for ICV and corpus callosum volumes, this suggests larger basal ganglia volumes in boys and with increasing age.

#### Sensitivity Analyses

Overall, there was good segmentation consistency across field strengths and antenatal maternal anaemia was associated with smaller total and regional brain volumes in infants between 3 and 24 months of age in both 3T and 64mT subsamples (Figure 4). However, there was more statistical power to detect significant findings in the larger ULF dataset. Given that maternal HIV has previously been associated with altered child brain structure,^41^ it was included in sensitivity analyses to ensure no confounding effects in the ULF subsample where group differences were observed. In sensitivity analyses across models, the effect of antenatal maternal anaemia on child brain structure remained largely unchanged and maternal HIV did not play a significant role in this subsample.

**Figure 4.**
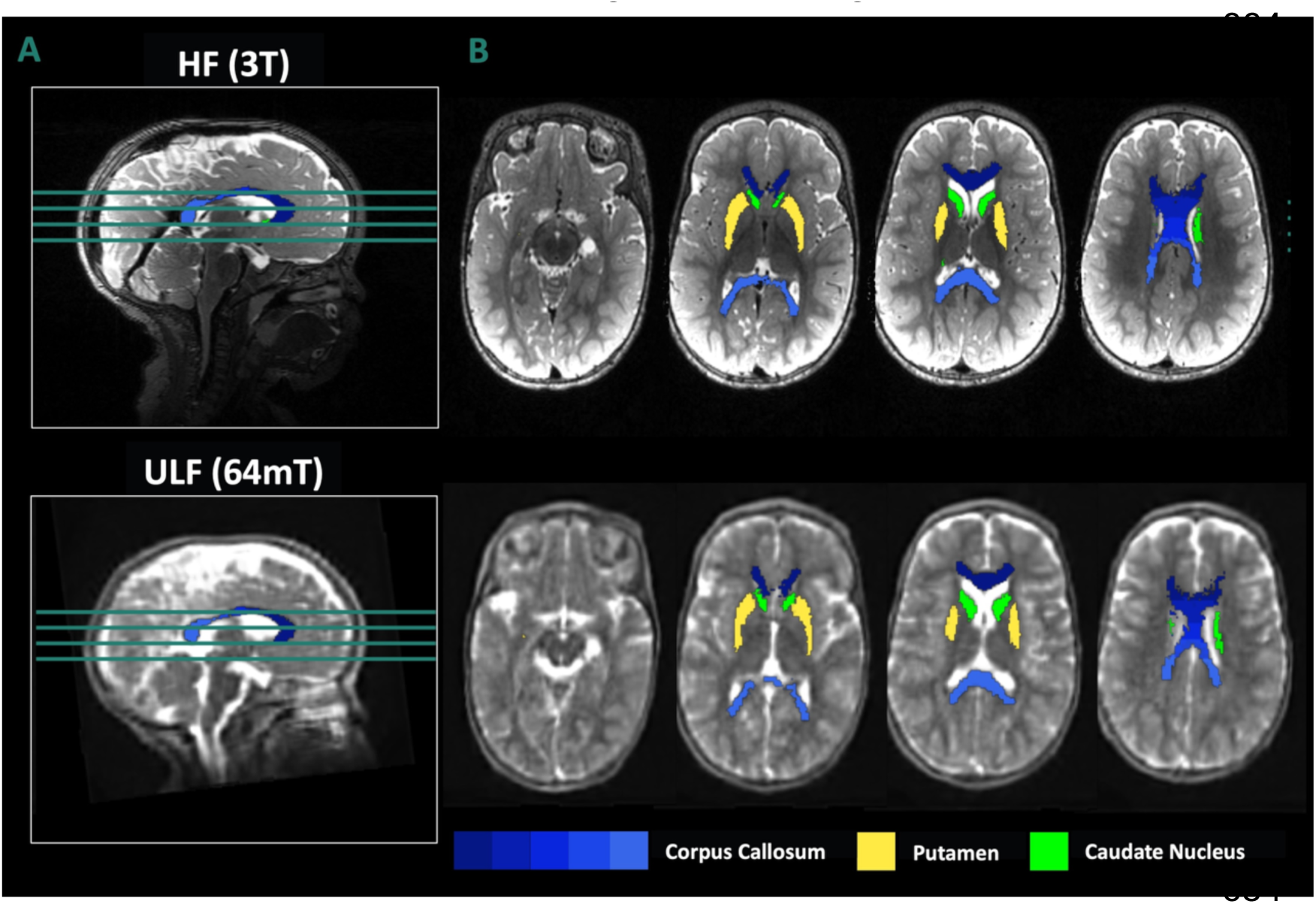
Segmentation Overlay of Brain Regions from Associated with Antenatal Maternal Anaemia in an Infant at 12 Months of Age Scanned using HF and ULF MRI A) Sagittal view of the brain demonstrating slices from inferior (bottom) to superior (top). B) Corresponding axial slices with segmentation overlay for brain regions of interest from inferior (left) to superior (right).

### Secondary Analysis: Postnatal Child Anaemia Status | ULF (64mT) MRI

#### Sample Characteristics: Postnatal Child Anaemia Status

In the ULF neuroimaging subsample (Supplementary Material; Table S2), 214 children had haemoglobin data from at least one study visit between 3-24 months. Of these children, 48.13% (103/214) were anaemic based on the lowest haemoglobin measurement recorded during this period. On average, the minimum postnatal haemoglobin measures from across scans for each infant (*n* = 214) was acquired around the 6-month visit (Mean [SD] = 7.11 [4.22] months, Range = 2.01 – 18.67 months). With repeated measures, 352 children had scans (average age at time of scan: 11.70 [5.54] months, age range of 2.66 – 25.49 months) with corresponding data on anaemia status based on the lowest haemoglobin measure between birth and the time of scan. Risk factors for child anaemia were identified (Supplementary Material; Table S2) as younger gestational age at birth (*p* = 0.034), and lower child anthropometry outcomes (birth length, *p* = 0.027). There were no other differences in maternal characteristics or infant characteristics between groups based on child anaemia status.

#### Modelling: Postnatal Child Anaemia Status

As an exploratory step, absolute brain volumes for ICV and ROIs were plotted against age with separate curves for children with anaemia and non-anaemic children in the repeated measures subsample (*n* = 352; Figure 5). No prominent group differences were observed on visual inspection. This was confirmed by LME models (child age or log of age, child sex, child anaemia status as predictors of interest) demonstrating no associations between child anaemia and child brain volumes for ICV (β = -0.03, *p* = 0.698) or the corpus callosum (β = - 0.15, *p* = 0.115), caudate nucleus (β = -0.13, *p* = 0.199), and putamen (β = -0.02, *p* = 0.744). Following on from this, the final antenatal maternal anaemia models established above (Primary Analysis) were replicated with the additional inclusion of child anaemia status as a covariate (*n* = 289). While the effects of antenatal maternal anaemia status remained similar, postnatal child anaemia was not a significant predictor of ICV (β = -0.04, *p* = 0.572), corpus callosum (β = -0.16, *p* = 0.134), caudate nucleus (β = -0.21, *p* = 0.059), or putamen (β = - 0.01, *p* = 0.942) volumes. Lastly, in exploring the additive effects of maternal and child anaemia (*n* = 92), it was found that anaemic children exposed to maternal anaemia in pregnancy did not have smaller ICV, corpus callosum, caudate nucleus, or putamen volumes than non-anaemic children exposed to maternal anaemia in pregnancy. Therefore, the combination of antenatal maternal and postnatal child anaemia was not significantly worse for child brain structure than antenatal maternal anaemia alone in this cohort.

**Figure 5.**
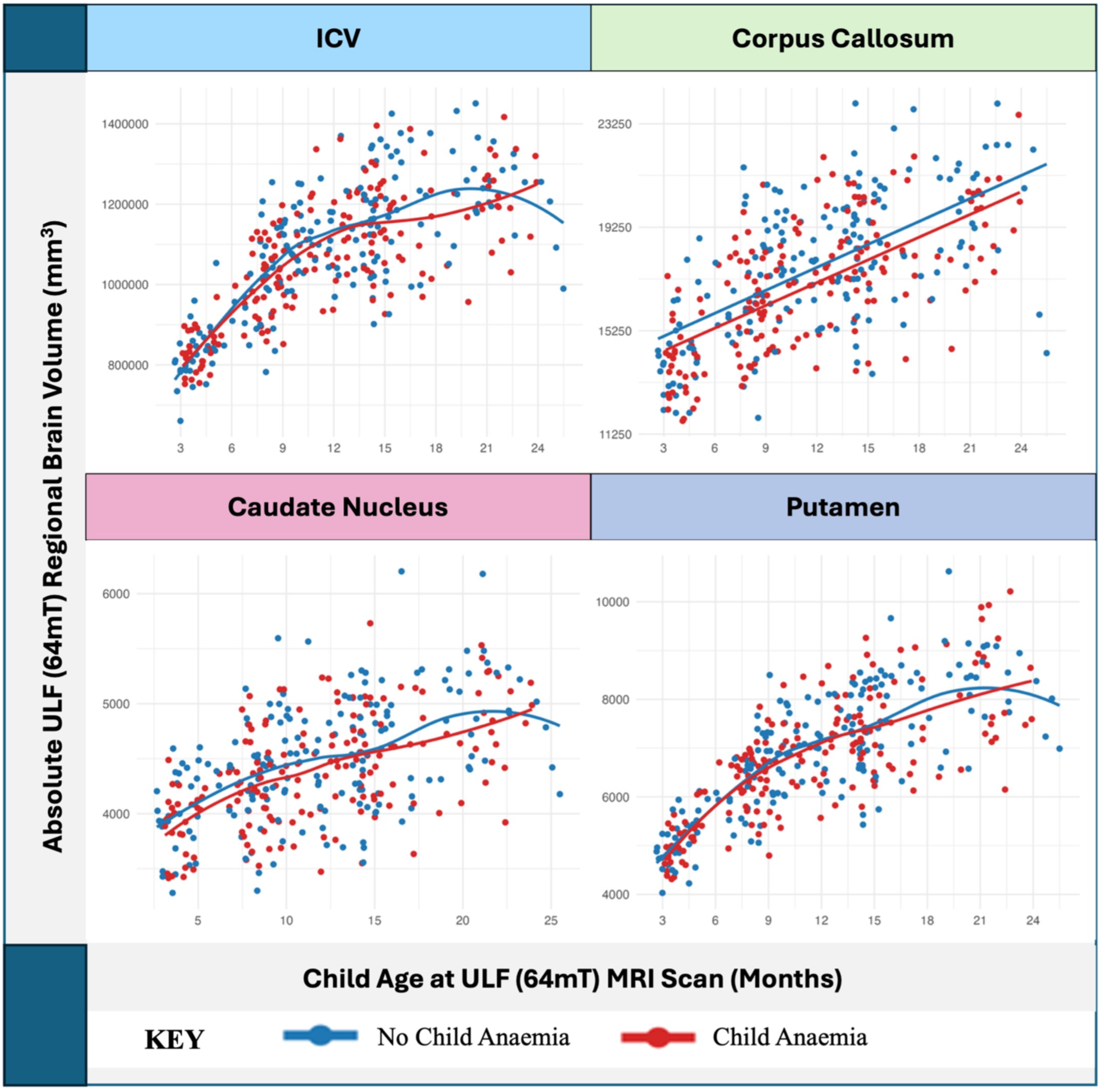
ULF (64mT; right) Absolute ICV and Regional Infant Brain Volumes By Age and Child Anaemia Status

### Tertiary Analysis: Iron Deficiency Status | ULF (64mT) MRI

Given that antenatal maternal anaemia (but not postnatal child anaemia) was significantly associated with ICV and regional child brain volumes, the role of iron deficiency was only investigated in mothers. This was based on the motivation for exploring the role of iron deficiency as a neurobiological mechanism underlying associations between anaemia and child brain structure.

#### Sample Characteristics: Antenatal Maternal Iron Deficiency Status

In the ULF infant neuroimaging subsample (Supplementary Materials; Table S3), 73 mothers (Mean [SD] age at enrolment: 28.80 [5.78] years, age range of 19.00 – 40.3 years) had adjusted serum ferritin data from pregnancy available. Minimum serum ferritin measures were predominantly acquired in the third trimester of pregnancy (67.12% [49/73], at a median (IQR) of 33 (30 – 35) weeks’ gestation. Of these mothers, 39.73% (29/73) were classified as iron deficient during pregnancy, after adjustment for inflammation. With repeated measures (*n* = 128), infants (average age at time of scan: 10.56 [5.32] months, age range of 2.69 – 25.07 months) born to mothers with iron deficiency in pregnancy had similar maternal and infant characteristics than infants born to mothers without iron deficiency in pregnancy.

#### Modelling: Antenatal Maternal Iron Deficiency Status

As an exploratory step, absolute brain volumes for ICV and ROIs were plotted against age (*n* = 128; Figure 6). Overall, no consistent group differences based on antenatal maternal iron deficiency status were observed. Based on LME models (child age or log of age, child sex, antenatal maternal iron deficiency as predictors of interest) assessing main effects, antenatal maternal iron deficiency was not associated with child brain volumes for ICV (β = -0.13, *p* = 0.317) or the corpus callosum (β = -0.02, *p* = 0.903), caudate nucleus (β = 0.06, *p* = 0.751), and putamen (β = 0.05, *p* = 0.730). The interactions between age and antenatal maternal iron deficiency status were also not predictive of ICV or regional brain volumes.

**Figure 6.**
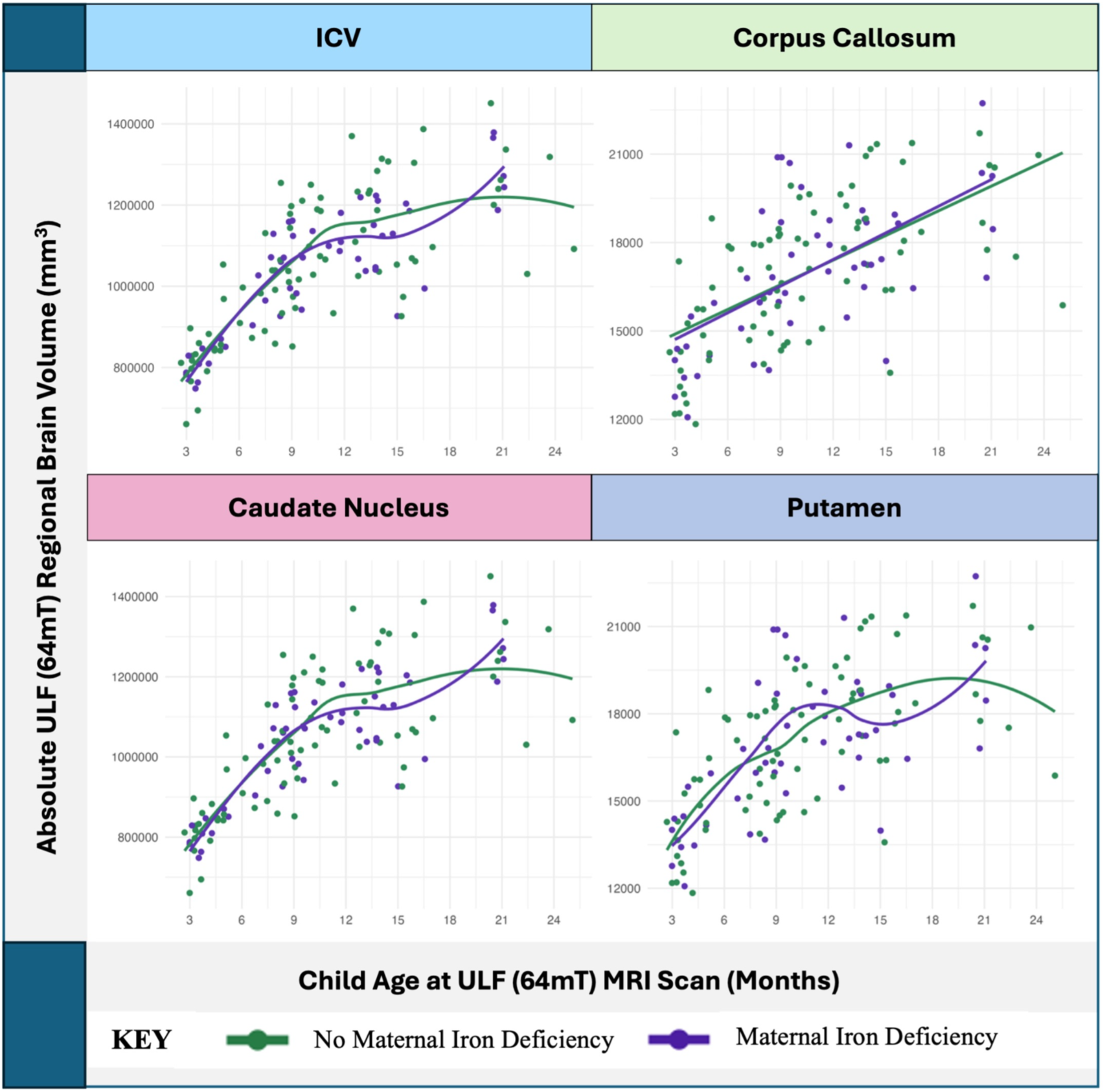
ULF (64mT; right) Absolute ICV and Regional Infant Brain Volumes By Age and Antenatal Maternal Iron Deficiency Status

## DISCUSSION

With the emerging recognition of the important role of antenatal maternal anaemia in altered neurodevelopment, there is a growing need to fully understand its impact across childhood. This neuroimaging work builds on previous South African research on toddlers^7^ and children,^21^ providing novel insight into the effects of anaemia in early infancy on brain structural growth. As the first study to explore these relationships in children under the age of two years, the results also suggest the age at which the effects of antenatal maternal anaemia may be most clearly detectable in the volumes of key subcortical structures. Overall, maternal anaemia in pregnancy, even when mild, was associated with smaller total (ICV) and subcortical (corpus callosum, caudate nucleus, and putamen) infant brain volumes (Figure 4), with group differences emerging at approximately 12 months of age. These effects were observed using both 3T and 64mT scanners, informing the feasibility of ULF MRI for ongoing research on anaemia in babies and young children from low-resource settings. Given the longitudinal nature of the cohort, these results inform current knowledge of regional brain volume trajectories in an LMIC context. With most brain growth curves being based on data from high-income countries,^42^ this is key for the valid interpretation of neuroimaging data on prevalent risk factors for child brain development across Africa.

Based on paediatric neuroimaging data from 3 – 24 months, exponential growth curves were observed for ICV as well as the caudate nucleus and putamen. This was characterised by rapid growth in early life with plateauing after 12 months of age. In contrast, the corpus callosum demonstrated steadier linear growth across the first two years of postnatal life. These patterns of structural brain development in South African babies from the Khula study are consistent with global brain charts across the lifespan^42^ and studies focusing on volumetric development in the first two years of childhood.^43–45^ However, as hypothesized, maternal anaemia in pregnancy was associated with reduced total brain volume (ICV) and smaller absolute volumes of the corpus callosum, caudate nucleus, and putamen.

Given the similarities in overall brain growth trajectories and observed volumetric differences by maternal anaemic status across MRI field strengths, this study suggests that the ULF system may provide sufficient sensitivity for detecting the effects of anaemia on the infant brain. This finding is key in validating the use of ULF MRI for both observational and experimental studies on the effects of risk factors and outcomes of interventions in ongoing anaemia research across the UNITY (Ultra- Low-Field Neuroimaging In The Young) network.^22^ This represents an important advancement toward increasing neuroimaging accessibility in LMICs faced with the highest burden of disease.

In the ULF subsample, infants born to mothers with anaemia in pregnancy had approximately 4% smaller total brain volumes and 3% smaller putamen volumes. These group differences became more prominent after 12 months, with a steeper deceleration in exponential growth at this age. Even larger effects were demonstrated for the caudate nucleus and corpus callosum, with an apparent interaction between antenatal maternal anaemia status and age across the first two years of life. Maternal anaemia in pregnancy was associated with approximately 4% smaller volumes of the caudate nucleus, with this effect becoming visually pronounced at 12 months (flatter exponential curve in exposed infants) and statistically significant for the first time at 18 months of age. Similarly, infants born to mothers who were anaemic during pregnancy had approximately 4% smaller corpus callosum volumes by 12 months and 7% smaller volumes by 24 months, with a slower rate of linear growth across the first two years of life in exposed infants. Overall, the impact of antenatal maternal anaemia differed in magnitude by age with effects emerging as infants approached 1 year of age.

These results from the Khula cohort are unique as they are the first to reveal the impact of antenatal maternal anaemia on child brain structure as early as infancy, when postnatal factors are less likely to have played as much of role. However, they are consistent with previously reported research on toddlers^7^ and children^21^ in a different South African cohort (DCHS), demonstrating remarkably comparable adjusted volume differences across age groups and studies for the caudate nucleus (4% smaller at 1.5 – 2 years in the Khula cohort, 5-6% smaller at 2 – 3 and 6 – 7 years in the DCHS cohort), corpus callosum (4% smaller at 1 year and 7% smaller at 2 years in the Khula cohort, 7 - 8% smaller at 2 - 3 and 6 - 7 years in the DCHS cohort), and putamen (3% smaller across first 2 years in the Khula cohort, 4% smaller at 2 – 3 years in the DCHS cohort). Taken together, these findings corroborate antenatal maternal anaemia as an important driver of altered child brain structure with regionally consistent effects emerging at 12 months and persisting throughout the first 6-7 years childhood. The replication of study results across these two representative cohorts also suggests that the demonstrated effects of anaemia on neurodevelopment may be generalisable across other communities in South Africa.

While this work reinforces the importance of pregnancy as the key period for optimised intervention, it also suggests the first year of life as a postnatal window during which brain structural differences in children exposed to maternal anaemia are not yet fully detectable. Based on previous research from the same cohort, the 12-month timepoint is also associated with an increased risk of postnatal iron deficiency as well as anaemia in infants born to anaemic mothers.^12^ It is possible that foetal iron loading^9,10^ and immuno-protective breastmilk with high iron bioavailability^46^ may serve as a buffer in the first year of life, reducing the risk of child iron deficiency anaemia and mitigating the effects of antenatal maternal anaemia on child brain structure during this period. This is relevant given its potential implications for the extension of targeted anaemia treatment strategies (e.g. iron supplementation) aimed at leveraging on protective factors in first 12 months of infancy. This may be particularly important for children at increased risk of anaemia themselves due to limited foetal iron loading in utero (antenatal maternal iron deficiency or pre-term birth)^47^ and postnatal factors such as poor nutrition or infection in early life.^12^

However, similar to what was observed in toddlers^7^ and school-age children^21^ from the DCHS, child anaemia was not associated with infant brain volume differences compared to those who were not anaemic in this Khula study. Furthermore, child anaemia did not moderate or worsen the effect of antenatal maternal anaemia on child brain volumes. While it is possible that child anaemia may still impact other aspects of brain development such as white matter maturation, volumetric outcomes seem to be largely driven by maternal anaemia in pregnancy. On this basis, the emergence of group differences at 12 months is unlikely to be caused by an interaction between antenatal and postnatal factors. Instead, the neurodevelopmental effects of anaemia in pregnancy may have been present earlier than 12 months, but masked by rapid overall brain growth and simultaneous asynchronous volume increases in other regions.^44,48^ With the plateauing and stabilisation of ICV and regional brain volumes at 12 months onwards, pre-existing patterns may have simply become clearer for disentanglement and detection. Therefore, while the first year of life presents an opportunity for further investigation and caution, we emphasize the importance of timing with the pregnancy period remaining critical for optimised outcomes.

Prevention and intervention strategies could be improved by targeting the underlying aetiologies of anaemia. Given that approximately 50% of anaemic cases can be attributed to iron deficiency in this cohort,^12^ this was investigated as an independent risk factor for altered child brain structure. However, the results of this study found no group differences in total or regional infant brain volumes based on maternal iron deficiency status in pregnancy. In explaining this finding, it is noted that the analysis on iron deficiency status may have been limited by insufficient power due to having half the sample size used for the anaemia analysis. While there are methodological limitations to our interpretation of this finding, the results do suggest that an absolute iron deficiency in mothers may not be the key neurobiological driver of observed associations between anaemia and regional infant brain volumes. Additionally, maternal anaemia was more common in mothers living with HIV, suggesting that “anaemia of chronic disease” may be the most relevant mechanism in this cohort. In these cases, chronic inflammation associated with infectious disease could be contributing to a functional iron deficiency characterised by sufficient iron stores but limited iron bioavailability.

Given that serum ferritin is an acute phase reactant that becomes transiently elevated in high inflammatory states, this study adjusted serum ferritin concentrations based on acute and chronic inflammatory biomarkers. Although this was necessary for the accurate classification of iron deficiency status, it does not preclude the possibility of iron stores not being accessible for use due to increased sequestration in the case of inflammation. If the effect of anaemia on child brain structure was being driven by a functional iron deficiency, comparing iron deficiency status based on absolute (albeit adjusted) serum ferritin values may not be appropriate. Future studies should focus on inflammatory mechanisms less sensitive to inflammation such as Transferrin Saturation (TSAT)^49–52^ and Reticulocyte Haemoglobin concentration (Ret-He)^53^ to make comparisons based on iron bioavailability.

In conclusion, antenatal maternal anaemia emerged as an important factor associated with reduced total brain volume as well as smaller corpus callosum, caudate nucleus, and putamen volumes. These findings were evident even with most cases of maternal anaemia being mild. This study is the first to demonstrate that the impact of maternal anaemia in pregnancy on child brain structure is evident as early as infancy, with effects detectable on brain volumes at 12 months of age. Although child anaemia was not associated with smaller total or regional brain volumes, the first year of life may present as an extended window for further intervention. Mothers of low household income living with HIV were at greatest risk of anaemia in pregnancy, suggesting that food insecurity due to poor socio-economic status and inflammation associated with infectious disease may be important factors for consideration. Antenatal maternal iron deficiency was not associated with smaller total or regional infant brain volumes, however, this analysis was limited by a smaller sample. Future research should include additional iron metrics for the assessment of iron bioavailability status in mothers with high levels of inflammation due to infectious disease, and intervention strategies should focus on the treatment of anaemia alongside appropriate infection management. Given the demonstrated validity of ULF for research on anaemia in infants and young children, this serves as a useful tool for ongoing research and trials in LMICs around the world.

## Data Availability

The de-identified data that support the findings of this study are available from authors upon reasonable request as per the Khula South Africa study guidelines.

## Acknowledgements

We would like to thank all the mothers and infants who participated in the Khula South Africa birth cohort study and made this research possible. We are also sincerely grateful to all the staff at both the Gugulethu Midwife Obstetric Unit as well as the University of Cape Town Neuroscience Institute who were involved in recruitment and data collection.

## ABBREVIATIONS

AGP: Alpha-1Acid Glycoprotein
CSF: Cerebrospinal Fluid
CUBIC: Cape Universities Body Imaging Centre
DCHS: Drakenstein Child Health Study
EMM: Estimated Marginal mean
Fe: Ferritin
Hb: Haemoglobin
hsCRP: highly sensitive C-Reactive Protein
HF MRI: High-Field Magnetic Resonance Imaging
HIV: Human Immunodeficiency Virus
ICV: Intracranial volume
LME: Linear Mixed Effects Model
LMIC: Low- and Middle-Income Country
MOU: Midwife Obstetrics Unit
MRI: Magnetic Resonance Imaging
MRR: Multi-Resolution Registration
PAE: Prenatal Alcohol Exposure
PTE: Prenatal Tobacco Exposure
RET-HE: Reticulocyte Haemoglobin
ROI: Region of Interest
SES: Socio-Economic Status
TSAT: Transferrin Saturation
ULF MRI: Ultra-Low-Field Magnetic Resonance Imaging
UNITY: Ultra-Low-Field Neuroimaging In The Young
QC: Quality Check
WHO: World Health Organization

## ARTICLE INFORMATION

### Author Contributions

*Conceptualization:* J.E. Ringshaw, M.R. Zieff, N.J. Bourke, S. Deoni, D.C. Alexander, D.K. Jones, S.C.R. Williams, K.A. Donald

*Data Curation:* J.E. Ringshaw, M.R. Zieff, N.J. Bourke, C. Casella, L.E. Bradford, J. O’Muircheartaigh

*Formal Analysis:* J.E. Ringshaw, M.R. Zieff

*Funding Acquisition:* J.E. Ringshaw, S.C.R. Williams, K.A. Donald

*Investigation:* L.E. Bradford, S.R. Williams, M. Miles, *Khula South Africa Data Collection

*Methodology:* J.E. Ringshaw, M.R. Zieff, N.J. Bourke, C. Casella, J. O’Muircheartaigh, S.C.R. Williams, K.A. Donald

*Project Administration:* M.R. Zieff, D. Herr, M. Miles, C. Bennallick

*Resources:* S.C.R. Williams, S. Deoni, K.A. Donald *Software:* N.J. Bourke, C. Casella, J. O’Muircheartaigh *Supervision:* D.J. Stein, S.C.R. Williams, K.A. Donald

*Validation:* J.E. Ringshaw, M.R. Zieff, N.J. Bourke, C. Casella, L.E. Bradford, S.R. Williams, J. O’Muircheartaigh, D.J. Stein, D.C. Alexander, D.K. Jones, S.C.R. Williams, K.A. Donald

*Visualisation:* J.E. Ringshaw

*Writing – Original Draft:* J.E. Ringshaw

*Writing – Review and Editing:* All authors.

*Khula South Africa Data Collection Team: Lauren Davel, Tracy Pan, Tembeka Mhlakwaphalwa, Bokang Methola, Khanyisa Nkubungu, Candice Knipe, Zamazimba Madi, Nwabisa Mlandu, Ringie Gulwa, Pamela Madikane

## Ethical Considerations

The Khula South Africa birth cohort study received ethical approval from the University of Cape Town Human Ethics Research Committee (HREC; 666/2021; 782/2022). All mothers provided written informed consent for cohort participation at Khula enrolment, and for their children to participate in study-specific procedures across timepoints. Given the longitudinal nature of the cohort, study consent is obtained from mothers annually for this cohort.

## Competing Interests

The authors report no competing interests.

## Funding

The Khula birth cohort was supported by the Wellcome Leap 1kD programme (The First 1000 Days; 222076/Z/20/Z). J.E. Ringshaw is supported by a Wellcome Trust International Training Fellowship (224287/Z/21/Z). The anemia analyses were also funded by the Gates Foundation (INV-023509) awarded to K.A. Donald. S.C.R. Williams is supported by the Bill and Melinda Gates Foundation (INV-047888) and the National Institute for Health and Care Research (NIHR) Maudsley Biomedical Research Centre (BRC). D.J. Stein is supported by the South African Medical Research Council (SAMRC). The funders had no role in study design, data collection and analysis, decision to publish, or preparation of the manuscript. For the purpose of open access, the authors have applied a CC BY public copyright licence to any Author Accepted Manuscript version arising from this submission.

## Notes

### Competing Interest Statement

The authors have declared no competing interest.

