## Supplementary Material for "Antenatal Maternal Anaemia and Infant Brain Structure: 3T and 64mT MRI Findings from South Africa"

---

**Table S1.** Zero-Order Correlations Between Potential Confounders, Antenatal Maternal Anaemia Status, and Brain Volumes

**Table S2.** Maternal and Infant ULF (64mT) Sample Characteristics According to Postnatal Child Anaemia Status

**Table S3.** Maternal and Infant ULF (64mT) Sample Characteristics According to BRINDA-Adjusted Antenatal Maternal Iron Deficiency Status

**Table S1.** Zero-Order Correlations Between Potential Confounders, Antenatal Maternal Anaemia Status, and Brain Volumes

| Correlation Matrix *r (p)* | | | | | | | |
|---|---|---|---|---|---|---|---|
| **HF (64mT) Subsample (*n* = 195)** | | | | | | | |
| **Variable[a]** | **Household Income** | **Maternal HIV** | **Maternal Anaemia** | **ICV** | **Corpus Callosum** | **Caudate Nucleus** | **Putamen** |
| **Household Income** | - | -0.278 (<0.001)*** | -0.127 (0.087) | -0.099 (0.182) | -0.003 (0.968) | -0.03 (0.690) | -0.10 (0.176) |
| **Maternal HIV** | | - | 0.103 (0.151) | 0.002 (0.983) | -0.026 (0.716) | -0.029 (0.683) | -0.008 (0.914) |
| **ULF (64mT) Dataset (*n* = 341)** | | | | | | | |
| **Household Income** | - | -0.241 (<0.001)*** | -0.189 (0.001) | 0.055 (0.333) | 0.019 (0.737) | 0.105 (0.066) | 0.058 (0.309) |
| **Maternal HIV** | | - | 0.131 (0.015)* | 0.028 (0.605) | 0.031 (0.566) | -0.029 (0.589) | -0.028 (0.610) |

[a] Covariates with observed differences in frequency between mothers with anaemia and mothers without anaemia were considered as potential confounders. Their inclusion in models assessing associations between antenatal maternal anaemia status and regional child brain volumes was considered based on zero-order correlations.
**p* is significant at <0.05, ***p* is significant at <0.01, ****p* is significant at <0.001.

**Table S2.** Maternal and Infant ULF (64mT) Sample Characteristics According to Postnatal Child Anaemia Status

| Variable[a] | Total Subsample ($n$ = 214)[a] | | |
|---|---|---|---|
| | **Child Anaemia[b] ($n$ = 103)** | **No Child Anaemia[b] ($n$ = 111)** | ***p*** |
| **Maternal Characteristics** | | | |
| Monthly household income (ZAR) [e,f] | | | |
| <1000 | 15 (14.56) | 22 (19.82) | 0.430 |
| 1000-5000 | 52 (50.49) | 47 (42.34) | |
| 5000-10000 | 21 (20.39) | 30 (27.03) | |
| >10000 | 4 (3.88) | 3 (2.70) | |
| Education[e] | | | |
| Primary | 1 (0.97) | 4 (3.60) | 0.580 |
| Some secondary | 53 (51.46) | 53 (47.75) | |
| Completed secondary | 39 (37.86) | 38 (34.23) | |
| Some Tertiary | 6 (5.83) | 11 (9.91) | |
| Completed Tertiary | 4 (3.88) | 5 (4.50) | |
| Employed[f] | 27 (26.21) | 39 (35.14) | 0.242 |
| Age Enrolment (years) | 28.78 (5.75)<br>[18.00 – 40.70] | 29.12 (5.85)<br>[19.00 – 42.60] | 0.671 |
| Anaemia during pregnancy[f] | 29 (28.16) | 27 (24.32) | 0.753 |
| Smoking during pregnancy[e] | 4 (3.88) | 6 (5.41) | 0.750 |
| Alcohol during pregnancy | 9 (8.74) | 4 (3.60) | 0.199 |
| Depression during pregnancy | 17 (16.50) | 22 (19.82) | 0.652 |
| HIV infection during pregnancy | 38 (36.89) | 39 (35.14) | 0.900 |
| **Infant Characteristics** | | | |
| Haemoglobin (g/dL)[g] | 9.96 (0.72)<br>[7.30 – 11.00] | 11.42 (0.79)<br>[9.50 – 14.1] | <0.001*** |
| Age at Hb Measurement (Months)[g] | 6.60 (4.09)<br>[2.33 – 18.67] | 7.40 (4.32)<br>[2.01 – 18.05] | 0.170 |
| Sex (boys) | 49 (47.57) | 58 (52.25) | 0.584 |
| Gestational age at birth (weeks)[d] | 38.80 (2.17)<br>[30.00 – 42.00] | 39.40 (1.35)<br>[35.00 – 42.00] | 0.034* |
| Child age at scan (months)[h] | 11.50 (5.51)<br>[3.12 – 23.92] | 11.91 (5.58)<br>[2.66 – 25.49] | 0.487 |
| HIV Infection[e] | 1 (0.97) | 0 (0.00) | 0.463 |
| Birth weight (g)[c,d] | 3132.21 (559.55)<br>[1180.00 – 4390.00] | 3164.91 (440.02)<br>[2160.00 – 4750.00] | 0.639 |
| Birth length (cm)[c,d] | 49.31 (4.01)<br>[33.00 – 57.00] | 50381 (2.81)<br>[41.00 – 57.00] | 0.027* |
| Birth head circumference (cm)[c] | 34.33 (1.81)<br>[28.00 – 38.00] | 34.71 (1.54)<br>[31.00 – 40.00] | 0.106 |

Abbreviations: Hb, haemoglobin; HIV, Human Immunodeficiency Virus; g, grams; cm, centimetres; ZAR, South African Rand.

SI conversion factor: To convert to haemoglobin grams per litre, multiply by 10.

[a] The total and group subsamples represent the number of unique subjects, excluding repeated measures data for children with multiple scans, to avoid duplication of observations for infants with multiple scans. Values for continuous variables are presented as: mean (standard deviation) [range]. Values for categorical variables are presented as: count (%).

[b] Dichotomous classifications for postnatal child anaemia at each study visit were determined using age-specific cut-offs for haemoglobin based on WHO guidelines for children over 6 months and local guidelines for children under 6 months. An overall classification for child anaemia status was determined based on the diagnosis of anaemia at least once across study visits. This was used for the assessment of group differences in sample characteristics between infants who had ever been anaemic versus infants who had never been anaemic between 3 and 24 months of age.

[c] The birth anthropometry were conducted by trained labour staff in the ward. Infant length was measures in cm to the nearest completed 0.5cm and weight was measured in kgs.

[d] Levene's test was significant. T-test results were interpreted based on equal variance not assumed

[e] Fisher's exact test result interpreted due to one or more cells having an expected count of less than 5.

[f] Missing Values: Maternal employment ($n$ = 2), monthly household income ($n$ = 20), infant HIV infection ($n$ = 147), maternal anaemia status ($n$ = 37)

[g] Minimum child haemoglobin and corresponding age were determined for each unique infant ($n$ = 214) from data across study visits in the repeated measures subsample.

[h] Mean child age at scan was calculated for the full subsample of scans ($n$ = 341) including repeated measures, given that each child may have had multiple scans at different ages from different study visits. The group summary statistics for child age at scan represent the mean, standard deviation, and range for each independent observation.

**p* is significant at <0.05, ***p* is significant at <0.01, ****p* is significant at <0.001.

**Table S3. Maternal and Infant ULF (64mT) Sample Characteristics According to BRINDA-Adjusted Antenatal Maternal Iron Deficiency Status**

| Variable[a] | Total Subsample (*n* = 73)[a] | | |
|---|---|---|---|
| | Maternal Iron Deficiency [b] (*n* = 29) | No Maternal Iron Deficiency [b] (*n* = 44) | *p* |
| **Maternal Characteristics** | | | |
| Adjusted serum ferritin (µg/L) [d] | 11.02 (2.40) [6.25 – 4.81] | 35.09 (23.36) [15.03 – 139.28] | <0.001*** |
| Gestational age at serum ferritin measurement [f] | 33.86 (3.31) [28.00 – 40.00] | 31.81 (3.31) [27.00 – 38.00] | 0.032* |
| Pregnancy trimester serum ferritin measured [e,f] | | | |
| First | 2 (6.90) | 6 (13.64) | 0.112 |
| Second | 4 (13.79) | 13 (29.55) | |
| Third | 15 (51.72) | 13 (29.55) | |
| Trimester unknown | 8 (27.59) | 12 (27.27) | |
| Monthly household income (ZAR)[e] | | | |
| <1000 | 6 (20.69) | 8 (18.18) | 1 |
| 1000-5000 | 13 (44.83) | 19 (43.18) | |
| 5000-10000 | 5 (17.24) | 7 (15.90) | |
| >10000 | 0 (0.00) | 0 (0.00) | |
| Education[e] | | | |
| Primary | 0 (0.00) | 1 (2.27) | 0.485 |
| Some secondary | 13 (44.83) | 19 (43.18) | |
| Completed secondary | 15 (51.72) | 18 (40.91) | |
| Some tertiary | 0 (0.00) | 4 (9.09) | |
| Completed tertiary | 1 (3.45) | 2 (4.55) | |
| Employed | 10 (34.48) | 12 (27.27) | 0.692 |
| Age at enrolment (years) | 29.12 (5.59) [19.60 – 40.30] | 28.59 (5.96) [19.00 – 39.40] | 0.705 |
| Smoking during pregnancy[e] | 0 (0.00) | 1 (2.27) | 1 |
| Alcohol during pregnancy[e] | 0 (0.00) | 3 (6.82) | 0.272 |
| Depression during pregnancy | 8 (27.59) | 8 (18.18) | 0.508 |
| HIV infection during pregnancy | 14 (48.28) | 15 (34.09) | 0.333 |
| **Infant Characteristics** | | | |
| Sex (male) | 19 (65.52) | 17 (38.64) | 0.045* |
| Gestational age at birth (weeks)[d] | 39.62 (1.02) [37.00 – 41.00] | 38.69 (1.99) [33.00 – 41.00] | 0.054 |
| Child age at scan (months, *n* = 128) [g] | 10.69 (5.09) [2.99 – 21.09] | 10.48 (5.49) [2.69 – 25.07] | 0/776 |
| HIV infection [e] | 0 (0.00) | (0.00) | N/A |
| Birth weight (g)[c] | 3278.97 (448.66) [2400.00 – 4360.00] | 3104.19 (508.62) [1780.00 – 4100.00] | 0.139 |
| Birth length (cm) [c] | 50.52 (3.00) [44.00 – 57.00] | 49.86 (3.33) [41.00 – 57.00] | 0.395 |
| Birth head circumference (cm) [c,d] | 34.88 (1.31) [32.00 – 38.00] | 34.32 (1.79) [30.50 – 37.00] | 0.156 |

*Abbreviations.* µg/L, micrograms per litre ; HIV, Human Immunodeficiency Virus; g, grams; cm, centimetres; ZAR, South African Rand

[a] The total and group subsamples represent the number of unique subjects, excluding repeated measures data for children with multiple scans, to avoid duplication of observations for infants with multiple scans. Values for continuous variables are presented as: mean (standard deviation) [range]. Values for categorical variables are presented as: count (%).

[b] Serum ferritin concentrations were adjusted for inflammation using the BRINDA regression correction approach. Antenatal maternal iron deficiency classified as adjusted serum ferritin concentrations <15µg/L

[c] The birth anthropometric measurements were conducted by trained labour staff in the ward. Infant length and head circumference were measured in cm to the nearest completed 0.5cm and weight was measured in kgs.

[d] Levene's test was significant. *T*-test results were interpreted based on equal variance not assumed.

[e] Fisher's exact test result interpreted due to one or more cells having an expected count of less than 5.

[f] Missing values: Trimester serum ferritin measured (*n* = 20), household income (*n* = 15), infant HIV (*n* = 48),

[g] Mean child age at scan was calculated for the full subsample of scans (*n* = 128) including repeated measures, given that each child may have had multiple scans at different ages from different study visits. The group summary statistics for child age at scan represent the mean, standard deviation, and range for each independent observation.

**p* is significant at <0.05, ** *p* is significant at <0.01, ****p* is significant at <0.001.
